# Trends in Racial and Ethnic Disparities in Barriers to Timely Medical Care Among US Adults, 1999 to 2018

**DOI:** 10.1101/2022.02.07.22270599

**Authors:** César Caraballo, Chima D. Ndumele, Brita Roy, Yuan Lu, Carley Riley, Jeph Herrin, Harlan M. Krumholz

**Affiliations:** Center for Outcomes Research and Evaluation, Yale New Haven Hospital, New Haven, Connecticut; Section of Cardiovascular Medicine, Department of Internal Medicine, Yale School of Medicine, New Haven, Connecticut; Department of Health Policy and Management, Yale School of Public Health, New Haven, Connecticut; Section of General Internal Medicine, Department of Internal Medicine, Yale School of Medicine, New Haven, Connecticut; Department of Chronic Disease Epidemiology, Yale School of Public Health, New Haven, Connecticut; Department of Pediatrics, University of Cincinnati College of Medicine, Cincinnati, Ohio; Division of Critical Care Medicine, Cincinnati Children’s Hospital Medical Center, Cincinnati, Ohio

## Abstract

**Background:** Racial and ethnic disparities in delayed medical care due to reasons not related to cost of care remain understudied. We aimed to describe recent 20-year trends in such disparities.

**Methods:** The study used data from the National Health Interview Survey from 1999–2018 and included individuals ≥18 years of age. Respondents were categorized by their sex, income, and self-reported race and ethnicity. The main outcomes were trends in disparities in any of 5 specific barriers to timely medical care: inability to get through by phone, no appointment available soon enough, long waiting times, inconvenient doctor’s office/clinic hours, or lack of transportation.

**Results:** The study included 590,603 adults (mean age 46.0 [SE, 0.07] years; 51.9% female). Of these, 4.7% were Asian, 11.8% Black, 13.8% Latino/Hispanic, and 69.7% White. In 1999, the proportion reporting any of the barriers to timely medical care was 7.3% among Asian, 6.9% among Black, 7.9% among Latino/Hispanic, and 7.0% among White individuals (P >0.05 for each difference with White individuals). From 1999 to 2018, this proportion increased across all 4 race/ethnicity groups (P<0.001 for each), slightly increasing the disparities between groups. In 2018, compared with White individuals, the proportion reporting any barrier was 2.1 and 3.1 percentage points higher among Black and Latino/Hispanic individuals (P=0.03 and P=0.001, respectively). The racial/ethnic disparities increased mostly among males and were attenuated when stratified by income level.

**Conclusions:** From 1999 to 2018, barriers to timely medical care increased for all populations with increasing disparities between racial/ethnic groups.

## BACKGROUND

There are racial and ethnic disparities in access to health care in the United States^1-4^ despite national efforts to eliminate them.^5-7^ For example, compared with White individuals, Black and Latino individuals persistently had a higher prevalence of lack of health insurance and unmet medical needs due to cost from 1999 to 2018.^4^ Though there has been considerable policy agreement on the need to reduce barriers to coverage and affordability, the healthcare community has given less attention to barriers to health care access that are not directly related to the cost of care and that may disproportionally affect patients, including racial and ethnic minorities, with social risk factors.

Black and Latino individuals, and those with low income, are more likely to experience barriers to timely medical care that are not cost-related,^8-12^ including long waiting times, inconvenient office hours, and lack of transportation. However, it is not known how racial and ethnic disparities in such barriers have changed over the last decades at the national level—and whether there has been any progress in eliminating them.

Accordingly, to comprehensively assess the nation’s performance for these indicators over 2 decades, we used the National Health Interview Survey (NHIS) to describe trends in racial and ethnic disparities in barriers to timely medical care that were not related to cost from 1999 to 2018. Given that there are differences in these barriers by sex and income,^10,13,14^ we also stratified the main findings by sex and income level.

## METHODS

### Data Source

We used data from the annual NHIS from 1999 to 2018 obtained from the Integrated Public Use Microdata Series Health Surveys (https://nhis.ipums.org/).^15^ The NHIS is a series of annual cross-sectional national surveys that provide information on the health of the noninstitutionalized population of the United States. The sample design uses a multistage area probability design, which adjusts for nonresponse and further allows for a national representative sampling of households and individuals, including traditionally underrepresented groups.^16^ The survey consists of a questionnaire divided into 4 cores (Supplemental Methods). In this study, we used data from the Sample Adult Core files (which contain responses from a detailed questionnaire from a randomly selected adult in each household). The mean conditional response rate and final response rate (which accounts for household and family nonresponse) of the Sample Adult Core survey during the study period were 81% and 64.8%, respectively. The code used to analyze these data is publicly available at https://doi.org/10.3886/E159861V1. The Institutional Review Board at Yale University exempted the study from review.

### Study Population

We included individuals ≥18 years of age. We excluded those who had missing information on barriers to timely medical care. Due to small numbers, we also excluded those who identified as non-Hispanic Alaskan Native, non-Hispanic American Indian, or non-Hispanic with no primary race, or “other” race, selected (details in Results section).

### Demographic Variables

We classified participants into 4 racial/ethnic groups based on their self-reported primary race and ethnicity: non-Hispanic Asian, non-Hispanic Black/African American, Latino/Hispanic, and non-Hispanic White. From the data, we also obtained age, sex, geographic region (Northeast, North Central/Midwest, South, West), and self-reported household income level. Based on the household income level relative to the respective year’s Federal Poverty Level from the United States Census Bureau, income level was categorized as low (<200% of the Federal Poverty Level) or middle/high (≥200% of the Federal Poverty Level).^4,17,18^

### Barriers to Timely Medical Care Not Related to Cost

Consistent with previous research,^10^ we defined the presence of barriers to timely medical care not related to cost by responses of “Yes” to the following question: “There are many reasons people delay getting medical care. Have you delayed getting care for any of the following reasons in the past 12 months? 1) You couldn’t get an appointment soon enough; 2) The (clinic/doctor’s) office wasn’t open when you could get there; 3) You couldn’t get through on the telephone; 4) Once you get there, you have to wait too long to see the doctor; or 5) You didn’t have transportation.” We determined a participant to have any barrier as responding “Yes” to any of the 5 specific barriers, while also determining the presence of each of the 5 barriers separately.

### Statistical Analysis

We estimated the annual prevalence of each of the barriers to timely medical care access using multivariable logistic regression models adjusting for age and region (Supplemental Methods). We then subtracted the annual prevalence among White respondents from the annual prevalence among the other race/ethnicity groups for that year, calculating SEs for the differences. Using these annual prevalences and differences, we calculated trends over the study period by fitting weighted linear regression models. Each observation was weighted by the inverse square of the SE of the prevalence to account for the varying precision of each estimate over time. Separately, we used a z-test to test for an absolute 1999–2018 difference in each barrier prevalence within each race and ethnic group and the differences between groups.

We then separately stratified the analysis described above by sex and household income. Due to the high prevalence of missing income data from participants’ non-response, our income-stratified analysis was based on the National Center for Health Statistics recommendations for multiply imputed data analysis in NHIS (Supplemental Methods).^19^

We also used ordered logistic regression models to estimate the proportion of individuals with 0, 1, 2, 3, or ≥4 specific barriers over the years (Supplemental Methods).

For all analyses, a 2-sided P-value <0.05 was used to determine statistical significance. All analyses were performed using Stata SE version 17.0 (StataCorp) and incorporated strata and weights to produce nationally representative estimates using the -*svy-* commands for structured survey data. All results are reported with 95% CIs. All person weights were pooled and divided by the number of years studied, following guidance from the NHIS.^20^

## RESULTS

### Study Population Characteristics

Of the 603,028 adults interviewed in the NHIS from 1999 to 2018, we excluded 5,752 who had missing information on barriers to timely medical care not related to cost. We also excluded 6,673 individuals from our analysis who identified as “other” race, did not identify with a primary race, or identified as Alaskan Native or American Indian (due to small numbers; Figure S1). The final sample was 590,603 adults with a mean (SE) age of 46.0 (0.07) years and of which 51.9% (95% CI: 51.7%, 52.0%) were female. Of these, 4.7% (95% CI: 4.5%, 4.8%) identified as Asian, 11.8% (95% CI: 11.5%, 12.1%) identified as Black, 13.8% (95% CI: 13.5%, 14.2%) identified as Latino/Hispanic, and 69.7% (95% CI: 69.3%, 70.2%) identified as White. Other characteristics of the population are described in Table 1 and Table S1.

**Table 1.**
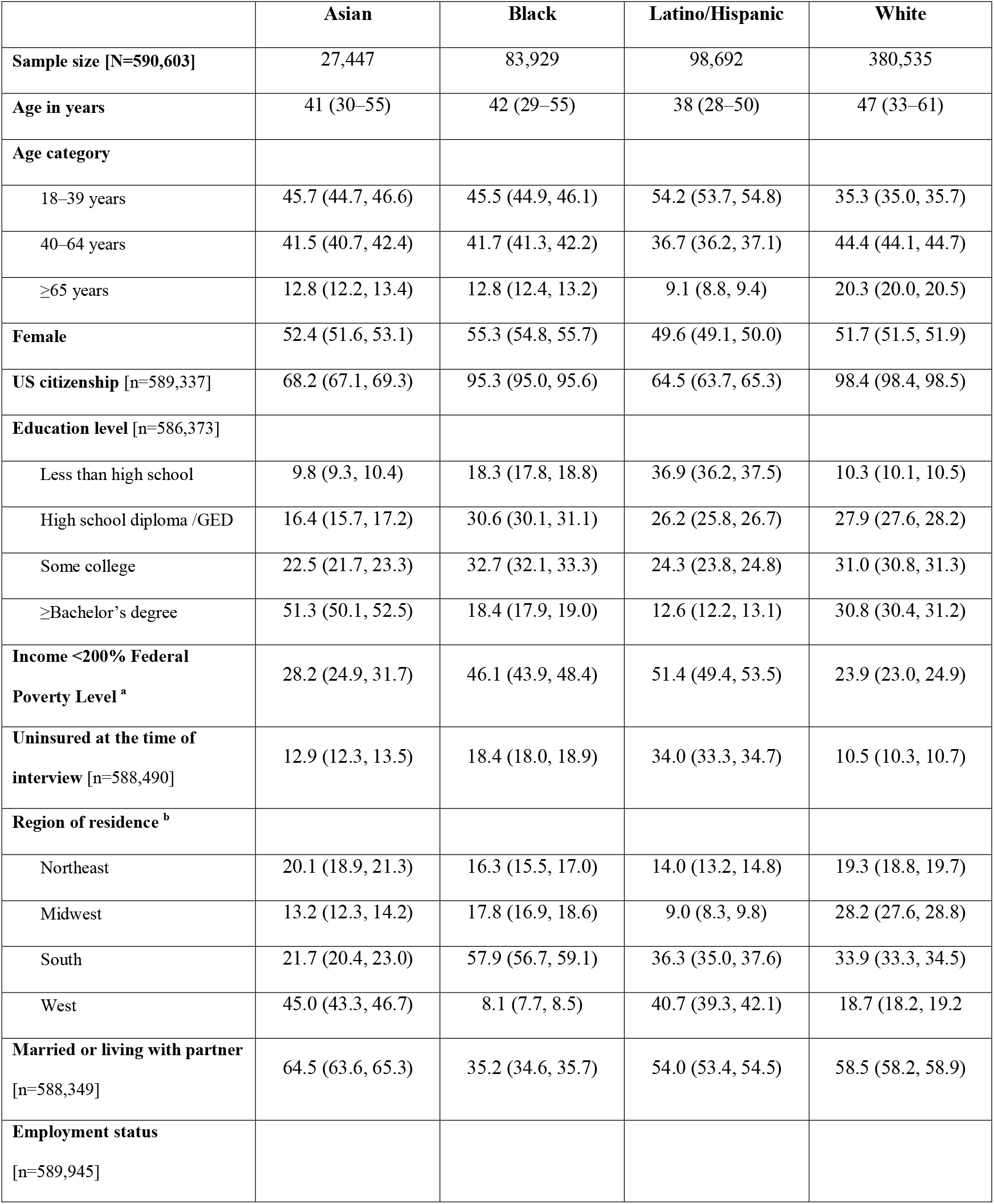
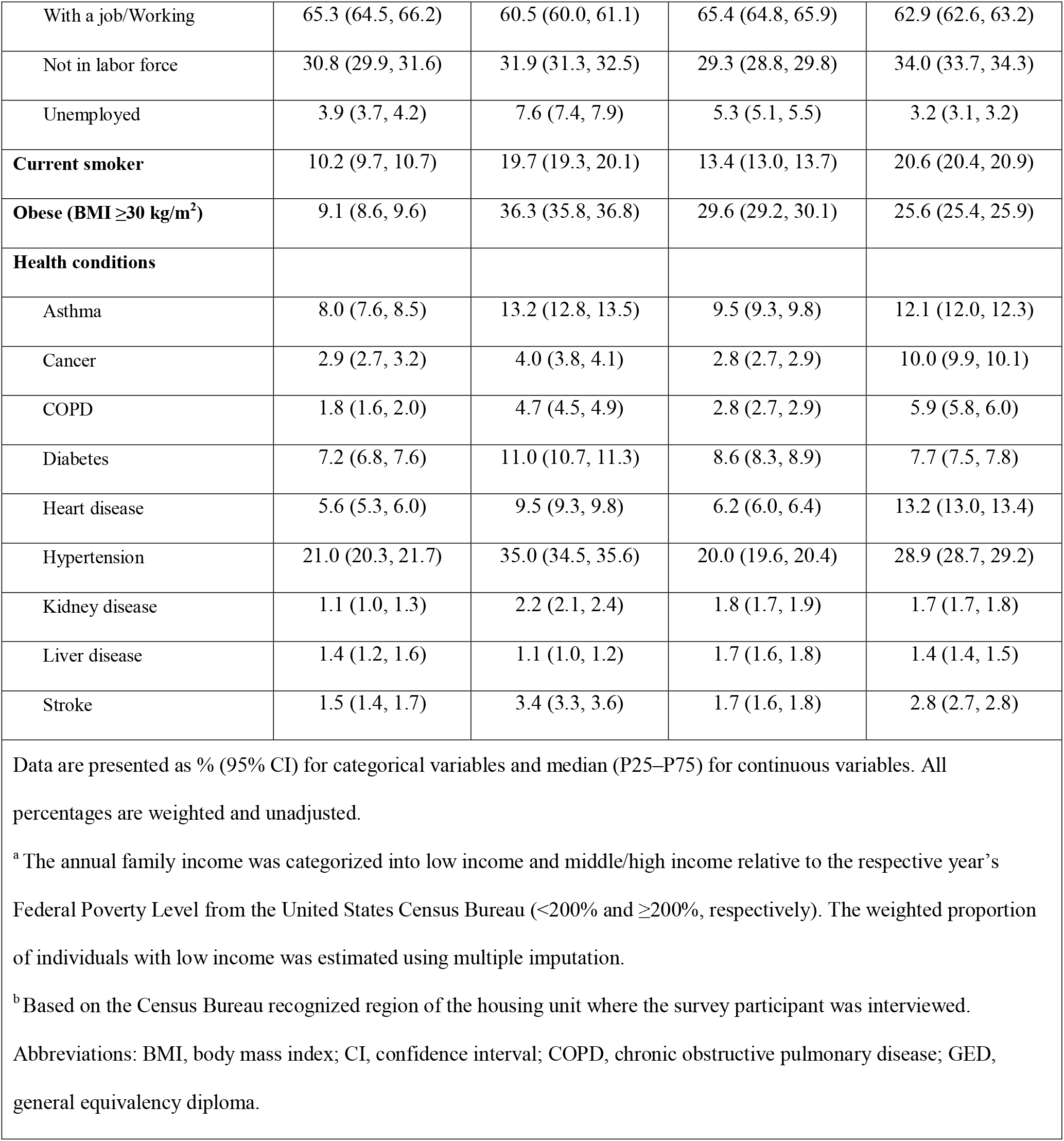
Study Population Characteristics.

### Temporal Trends in Racial and Ethnic Differences in Prevalence of Barriers to Timely Medical Care not Related to Cost

#### Any Barrier

In 1999, the overall proportion of individuals reporting any of the barriers to timely medical care was 7.1% (95% CI: 6.7, 7.4), and there were no significant differences between White people and Asian, Latino, and Black people (P=0.83, P=0.95, and P=0.12, respectively; Table 2). The adjusted estimated prevalence in 1999 was 7.3% among Asian individuals (95% CI: 5.5%, 9.5%), 6.9% among Black individuals (95% CI: 5.9%, 8.0%), 7.9% among Latino/Hispanic individuals (95% CI: 6.9%, 9.0%), and 7.0% among White individuals (95% CI: 6.6%, 7.5%) (Figure 1). From 1999 to 2018, prevalence increased in all 4 race/ethnicity groups (Table 2) (P<0.001 for each), slightly increasing the gap between White people and Black and Latino people. In 2018, the overall proportion reporting any barrier was 13.5% (95% CI: 12.8, 14.1). The proportion reporting any barrier, compared with the adjusted prevalence among White people (12.9% [95% CI: 12.3%, 13.6%]), was 2.1 percentage points higher among Black people and 3.1 percentage points higher among Latino/Hispanic people (P=0.03 and P=0.001, respectively). There was no significant difference in prevalence between Asian and White people (P=0.94). Similarly, Black and Latino/Hispanic people had the greatest prevalence of at least 1, 2, 3, or 4 barriers over the study period (Figure 2).

**Table 2.** Change in the Adjusted Prevalence of Barriers to Timely Medical Care Not Related to Cost from 1999–2018, by Race and Ethnicity.

|  | **Asian individuals** | **Black individuals** | **Latino/Hispanic individuals** | **White individuals** |
| --- | --- | --- | --- | --- |
|  | Percentage points (95% CI), p value | Percentage points (95% CI), p value | Percentage points (95% CI), p value | Percentage points (95% CI), p value |
| **Any barrier** |  |  |  |  |
| Change in prevalence, 1999–2018 | +5.74 (+2.38, +9.11), <0.001 | +7.95 (+5.94, +9.95), <0.001 | +8.07 (+6.03, +10.11), <0.001 | +5.85 (+5.06, +6.63), <0.001 |
| Change in difference with White individuals, 1999–2018 | -0.10 (-3.56, +3.35), 0.95 | +2.10 (-0.05, +4.25), 0.06 | +2.22 (+0.03, +4.40), 0.05 | - |
| Difference in 1999 | +0.22 (-1.82, +2.25), 0.83 | -0.03 (-1.15, +1.08), 0.95 | +0.88 (-0.23, +1.99), 0.12 | - |
| Difference in 2018 | +0.11 (-2.68, +2.90), 0.94 | +2.06 (+0.22, +3.91), 0.03 | +3.10 (+1.22, +4.98), 0.001 | - |
| **Specific barriers.** |  |  |  |  |
| **Individuals delayed care because** |  |  |  |  |
| **...they couldn't get through on the telephone** |  |  |  |  |
| Change in prevalence, 1999–2018 | +0.69 (-0.93, +2.32), 0.40 | +1.43 (+0.53, +2.33), 0.002 | +1.67 (+0.71, +2.64), <0.001 | +0.89 (+0.48, +1.29), <0.001 |
| Change in difference with White individuals, 1999–2018 | -0.19 (-1.87, +1.48), 0.82 | +0.54 (-0.45, +1.53), 0.29 | +0.79 (-0.26, +1.83), 0.14 | - |
| Difference in 1999 | -0.01 (-1.28, +1.27), 0.99 | -0.58 (-1.05, -0.11), 0.02 | -0.39 (-0.91, +0.12), 0.14 | - |
| Difference in 2018 | -0.20 (-1.29, +0.89), 0.72 | -0.04 (-0.91, +0.82), 0.92 | +0.39 (-0.51, +1.30), 0.40 | - |
| **...they couldn't get an appointment soon enough** |  |  |  |  |
| Change in prevalence, 1999–2018 | +3.60 (+1.11, +6.09), 0.01 | +4.49 (+3.12, +5.86), <0.001 | +4.35 (+2.84, +5.85), <0.001 | +4.07 (+3.46, +4.68), <0.001 |
| Change in difference with White individuals, 1999–2018 | -0.47 (-3.03, +2.09), 0.72 | +0.42 (-1.08, +1.92), 0.58 | +0.27 (-1.35, +1.90), 0.74 | - |
| Difference in 1999 | 0.00 (-1.45, +1.45), 0.99 | -0.88 (-1.56, -0.20), 0.01 | +0.28 (-0.49, +1.05), 0.47 | - |
| Difference in 2018 | -0.47 (-2.58, +1.64), 0.66 | -0.46 (-1.80, +0.88), 0.50 | +0.55 (-0.88, +1.99), 0.45 | - |
| **...once they got there, they had to wait too long to see the doctor** |  |  |  |  |
| Change in prevalence, 1999–2018 | +2.60 (+0.98, +4.22), 0.002 | +2.73 (+1.43, +4.02), <0.001 | +3.86 (+2.42, +5.29), <0.001 | +1.23 (+0.80, +1.66), <0.001 |
| Change in difference with White individuals, 1999–2018 | +1.37 (-0.31, +3.05), 0.11 | +1.50 (+0.13, +2.86), 0.03 | +2.63 (+1.13, +4.12), <0.001 | - |
| Difference in 1999 | -0.59 (-1.64, +0.47), 0.27 | +0.63 (-0.07, +1.32), 0.08 | +1.34 (+0.61, +2.08), <0.001 | - |
| Difference in 2018 | +0.78 (-0.52, +2.09), 0.24 | +2.12 (+0.95, +3.30), <0.001 | +3.97 (+2.66, +5.27), <0.001 | - |
| **...the clinic/doctor's office wasn't open when they could get there** |  |  |  |  |
| Change in prevalence from 1999–2018 | +1.28 (-0.48, +3.03), 0.15 | +1.45 (+0.46, +2.43), 0.004 | +1.81 (+0.79, +2.83), <0.001 | +1.58 (+1.14, +2.03), <0.001 |
| Change in difference with White individuals, 1999–2018 | -0.31 (-2.12, +1.50), 0.74 | -0.14 (-1.22, +0.94), 0.80 | +0.22 (-0.89, +1.34), 0.69 | - |
| Difference in 1999 | -0.09 (-1.30, +1.12), 0.89 | -0.52 (-1.01, -0.03), 0.04 | -0.19 (-0.72, +0.35), 0.50 | - |
| Difference in 2018 | -0.40 (-1.74, +0.95), 0.57 | -0.66 (-1.62, +0.30), 0.18 | +0.04 (-0.93, +1.01), 0.94 | - |
| **...they didn't have transportation** |  |  |  |  |
| Change in prevalence from 1999–2018 | -0.32 (-2.77, +2.14), 0.80 | +3.09 (+1.05, +5.13), 0.003 | +1.25 (+0.17, +2.33), 0.02 | +0.83 (+0.50, +1.16), <0.001 |
| Change in difference with White individuals, 1999–2018 | -1.15 (-3.63, +1.33), 0.37 | +2.26 (+0.19, +4.33), 0.03 | +0.42 (-0.71, +1.55), 0.47 | - |
| Difference in 1999 | +1.21 (-0.73, +3.15), 0.22 | +1.51 (+0.58, +2.45), 0.001 | +0.45 (-0.20, +1.11), 0.17 | - |
| Difference in 2018 | +0.36 (-0.76, +1.49), 0.53 | +3.38 (+2.24, +4.51), <0.001 | +1.00 (+0.33, +1.68), 0.004 | - |
| Data source is the National Health Interview Survey from years 1999–2018. For change in prevalence and change in difference: a positive sign (+) means the prevalence of each indicator (or its difference with White people) increased and a negative sign (-) means it decreased. Estimates were adjusted by age and US region. For each difference in prevalence (1999 and 2018), the corresponding excess weighted number of individuals experiencing it is shown in Supplemental Table S2. Income- and sex-stratified estimates are also shown in the Supplement.<br>Abbreviations: CI, confidence interval |  |  |  |  |

**Figure 1.**
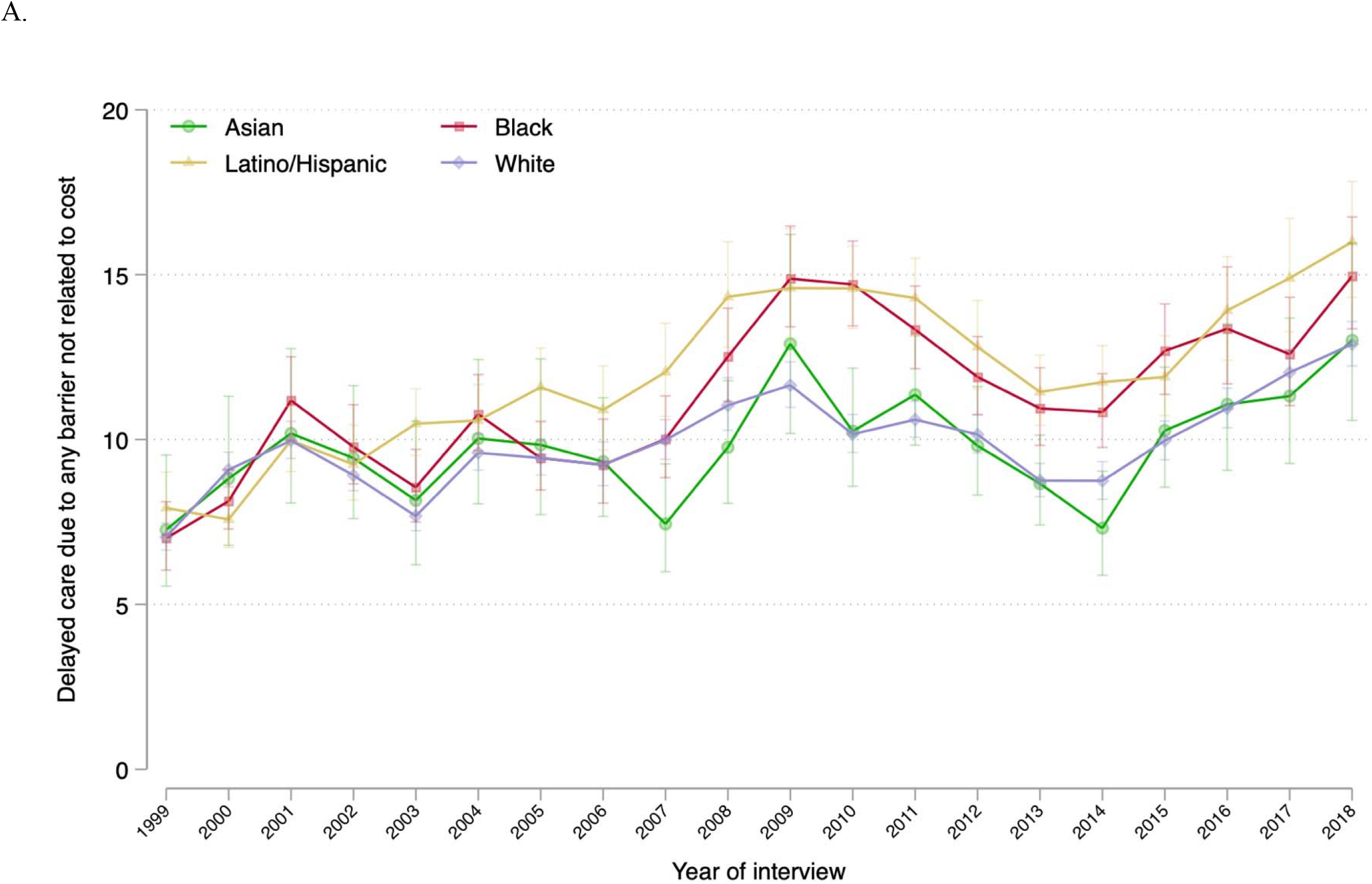
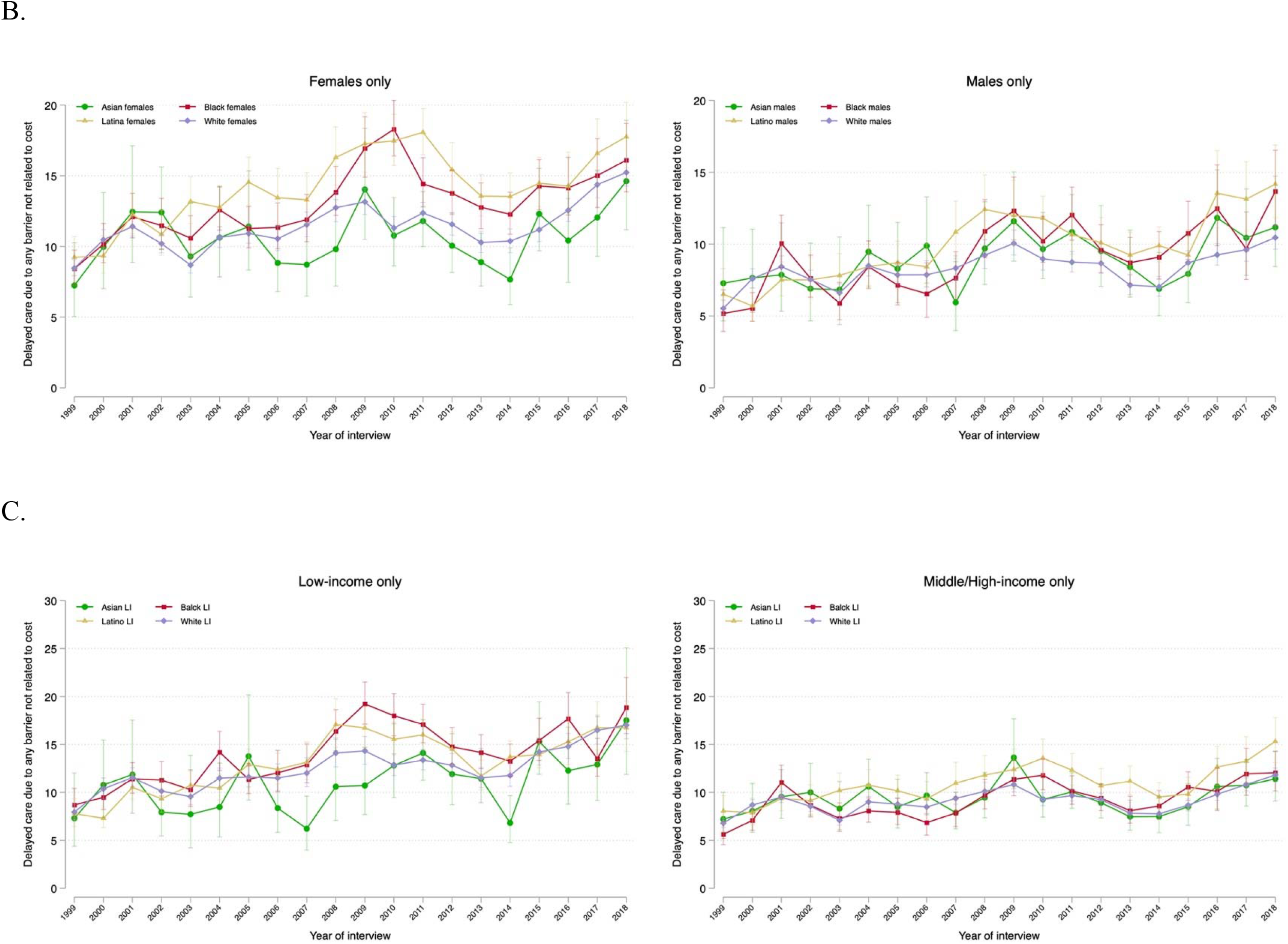
Trends in Annual Prevalence of Barriers to Timely Medical Care Among US Adults by Race and Ethnicity, 1999–2018. Data source is the National Health Interview Survey from years 1999–2018. Brackets represent the 95% confidence intervals. All estimates were adjusted by age and US region.

**Figure 2.**
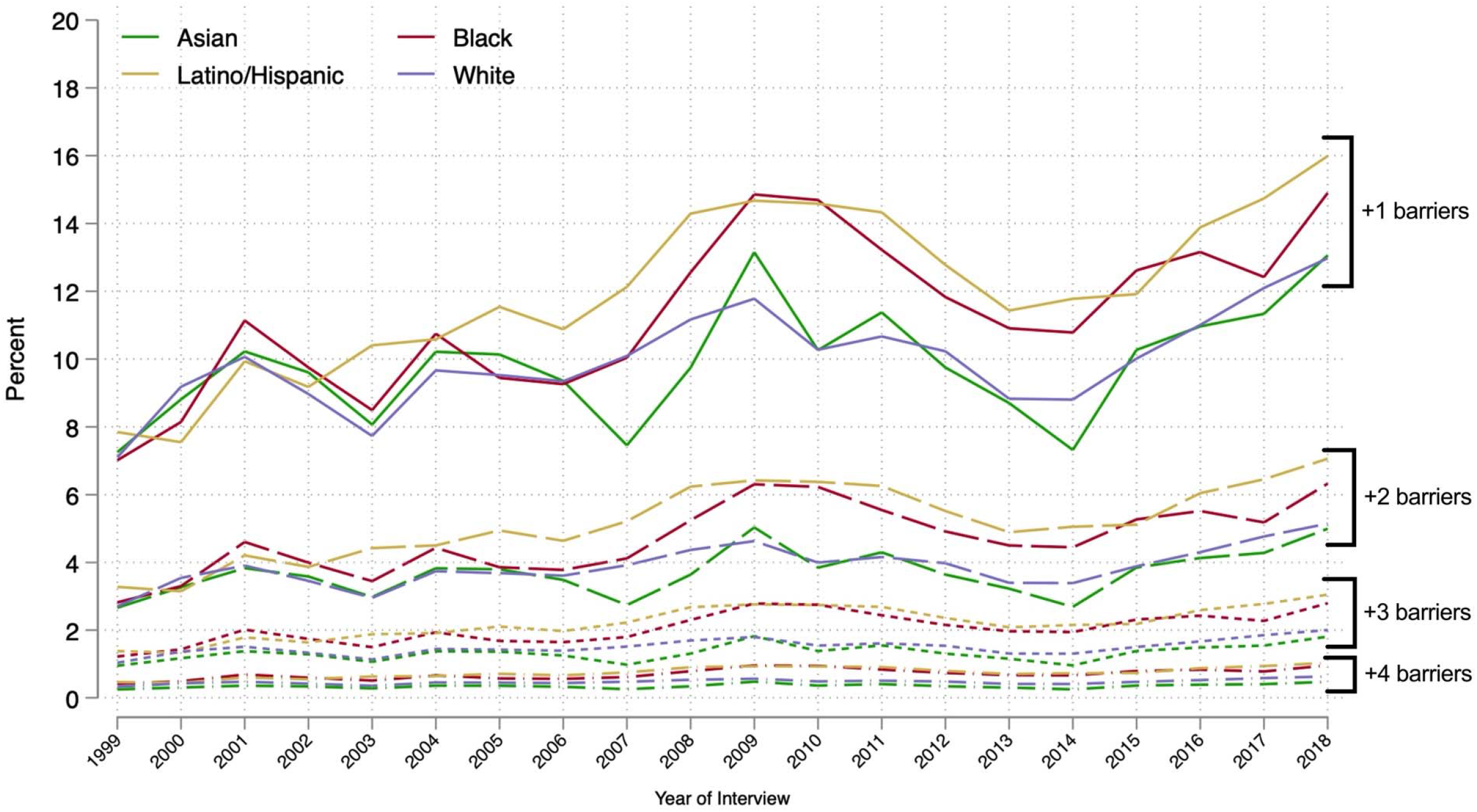
Trends in Estimated Ordered Number of Barriers to Timely Medical Care Not Related to Cost by Race and Ethnicity, 1999–2018. Data source is the National Health Interview Survey from 1999–2018. The ordered number of chronic conditions was estimated using ordered logistic regression. All estimates were adjusted by age and US region.

When estimates were stratified by sex, the prevalence of these barriers to timely medical care increased over time among both males and females. The racial/ethnic gap only increased among males, with no significant changes observed in the gap among females (Table S2). In 2018, compared with White males, the estimated prevalence was 3.2 percentage points higher among Black males and 3.7 points higher among Latino/Hispanic males (P=0.03 and P=0.01, respectively). When analyzed by income level, there were no significant changes in the differences between subgroups during the study period (Table S2) and the 1999 and 2018 differences between White and Black individuals were not significant within each income stratum. Among those with middle/high-income, the prevalence of barriers to timely medical care was 3.5 percentage points higher among Latino/Hispanic people compared with non-Hispanic White people (P=0.01; Table S2).

### Specific Barriers to Timely Medical Care

During the study period, each of the 5 barriers significantly increased in prevalence among Black, Latino/Hispanic, and White people (Figure 3 and Table 2). Among Asian people, the increase occurred only in the proportion of those who reported having delayed care due to long waiting times and because they could not get an appointment soon enough (+2.6 and +3.6 percentage points, respectively; P≤0.01 each).

**Figure 3.**
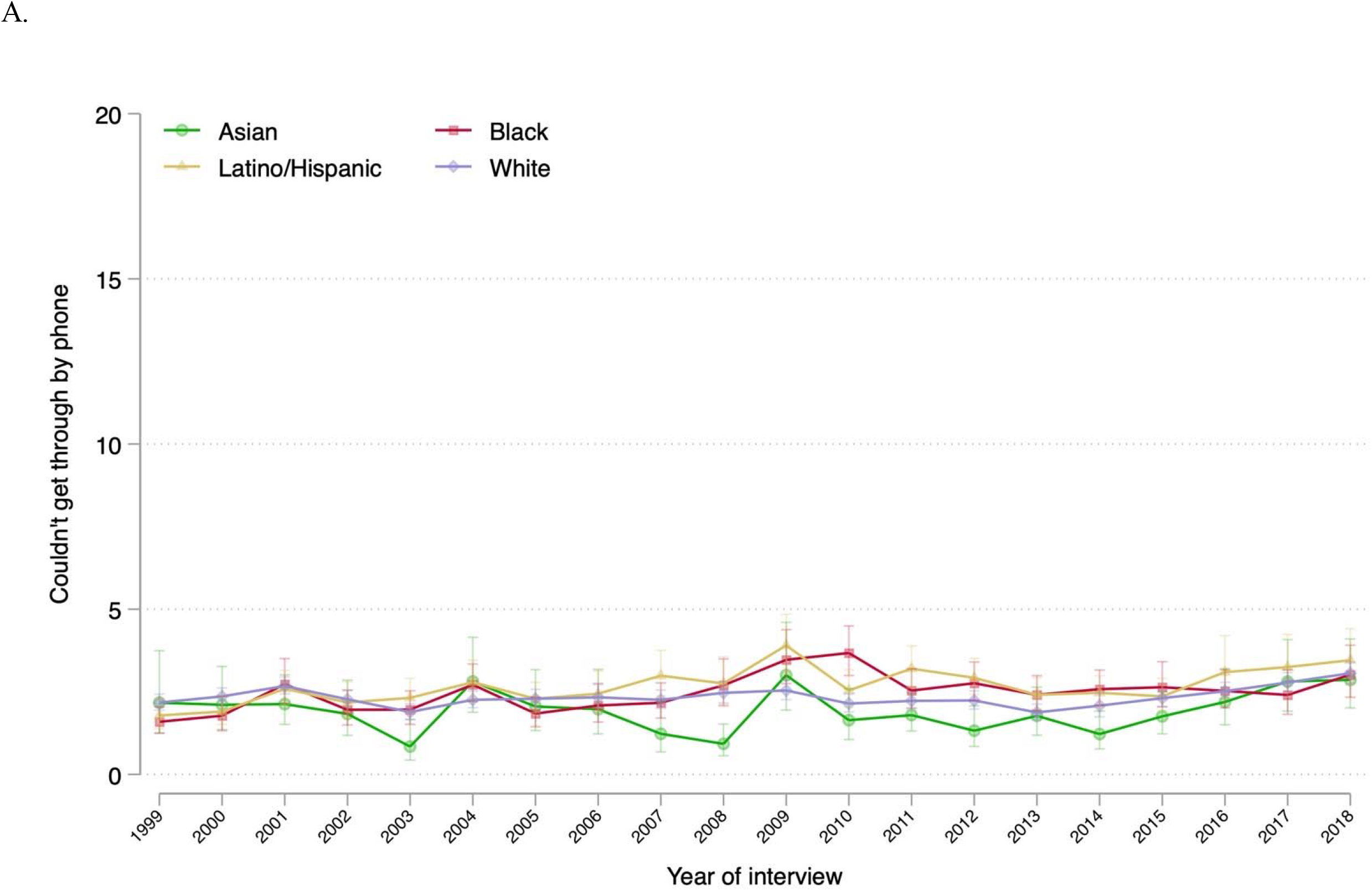
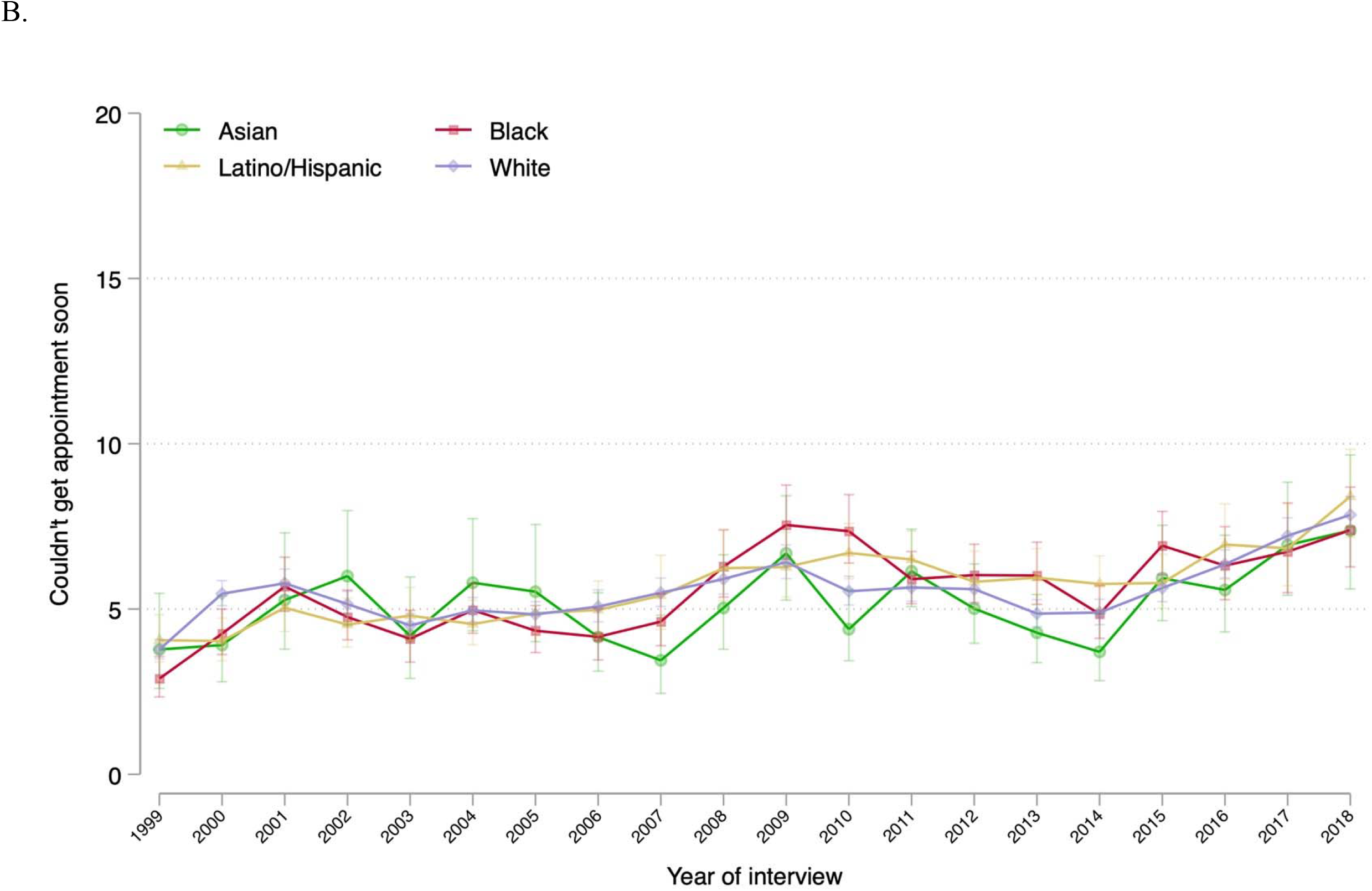
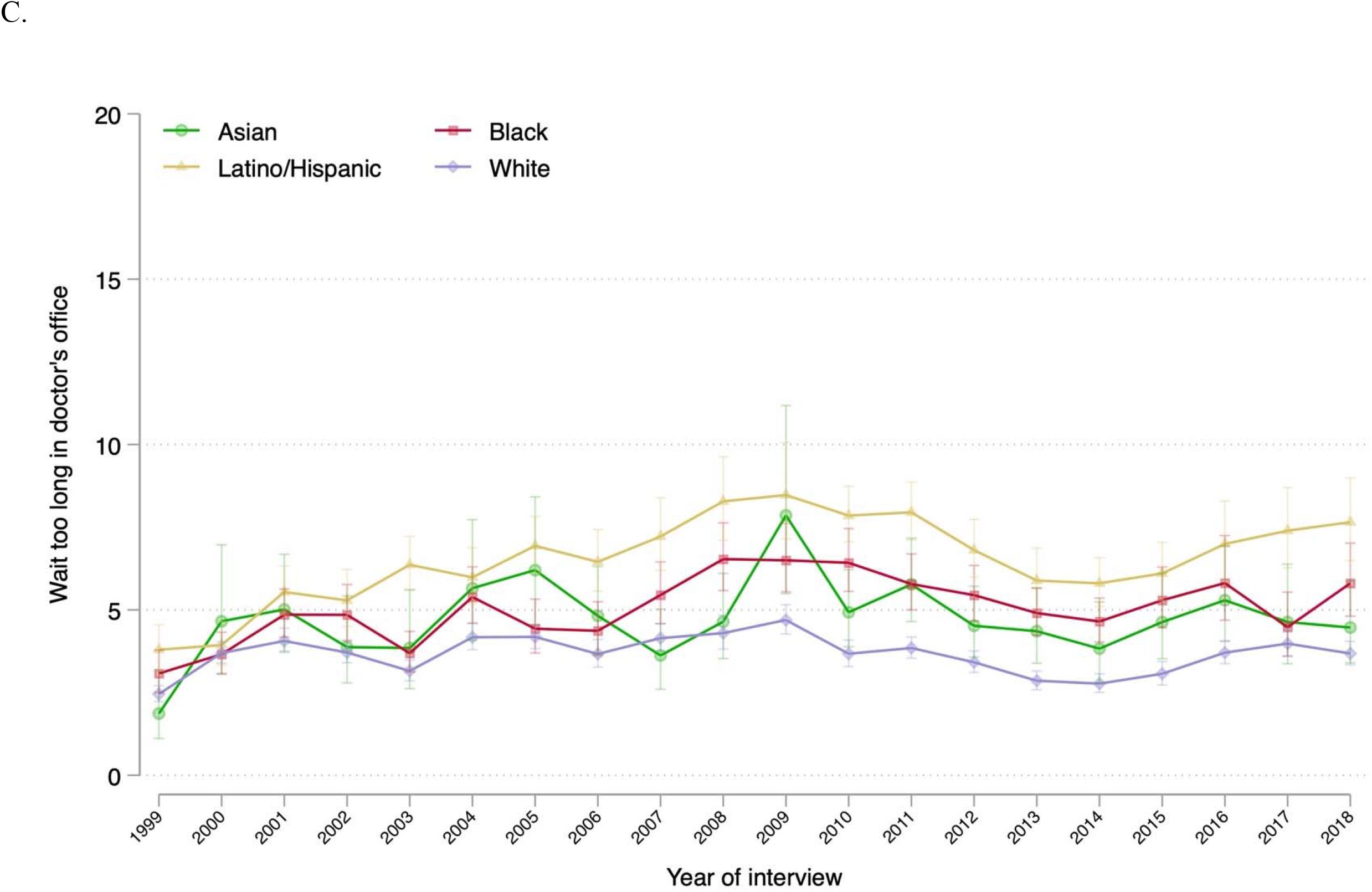
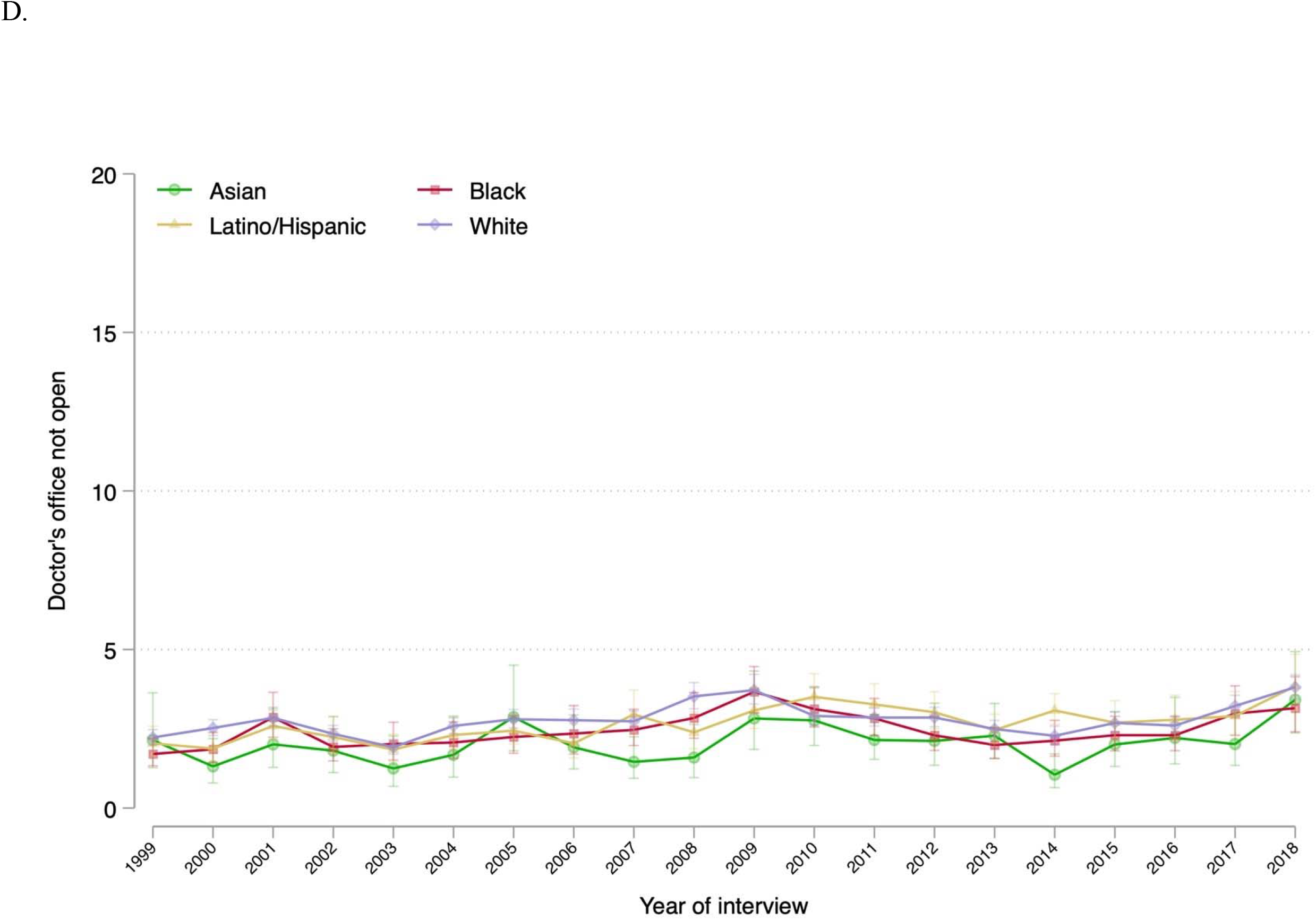
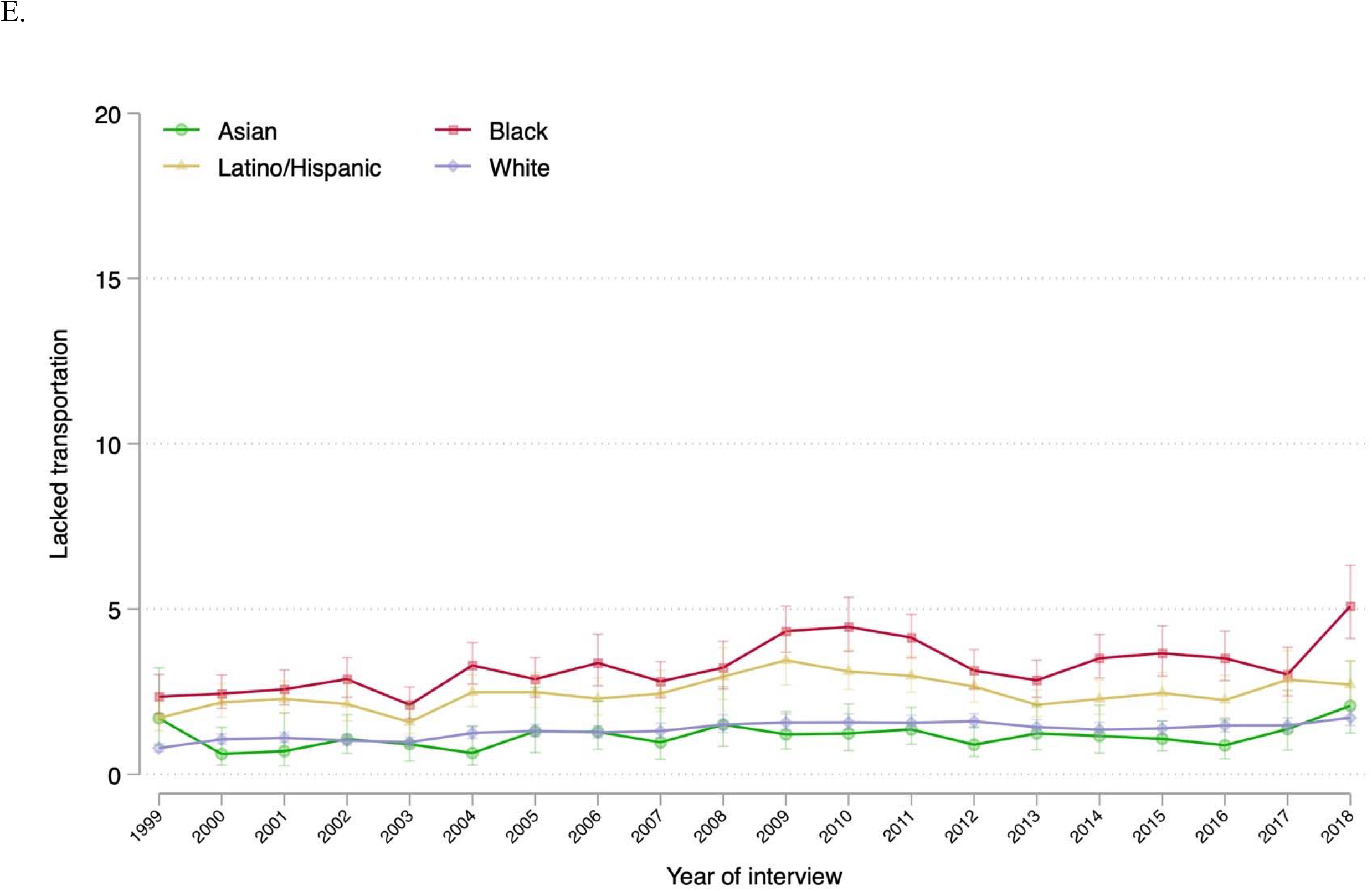
Trends in Annual Prevalence of Specific Barriers to Timely Medical Care Among US Adults by Race and Ethnicity, 1999–2018. Data source is the National Health Interview Survey from years 1999–2018. Brackets represent the 95% confidence intervals. All estimates were adjusted by age and US region.

There was a significant increase in the difference in prevalence between Black and White individuals who reported delaying care because of long waiting times at the clinic/doctor’s office and because of lack of transportation (increased by 1.5 percentage points and 2.3 percentage points, respectively [P=0.03 each]). In 2018, compared with the prevalence of each of these two barriers among White individuals (3.7% [95% CI: 3.3%, 4.1%] and 1.7% [95% CI: 1.5%, 2.0%], respectively), the prevalences were higher by 2.1 percentage points and 3.4 percentage points among Black individuals, respectively (P<0.01 for each). Such differences were still significant when stratified by sex and income level (Tables S4 and S7). However, among those with middle/high income, the difference between Black and White people who reported delaying care due to long waiting times was not significant in 2018 (Tables S4 and S7).

In addition, the difference in prevalence between Latino/Hispanic and White people who reported delaying care due to long waiting times widened significantly (increased by 2.6 percentage points, P<0.001). In 2018, compared with the prevalence among White individuals, the proportion of Latino/Hispanic individuals who experienced this barrier was 4.0 percentage points higher (P<0.001). This difference was still significant when stratified by sex and income level (Table S5). In the same year, the prevalence of Latino/Hispanic people who reported delayed care due to lack of transportation was 1.0 percentage points higher (Table 2) than that of White people, with the difference mainly among females and individuals of middle or high income (Table S7).

The change in prevalence difference between subgroups for the other 3 barriers from 1999–2018 was not significant (Table 2), with little variation by sex or household income level (Figures S2–S6 and Tables S2–S7).

## DISCUSSION

In this nationally representative study, we found that from 1999 to 2018, the overall estimated proportion of people who reported barriers to timely care nearly doubled, increasing from 7.1% to 13.5%, and the increase was not proportionate across the 4 racial and ethnic groups. During this period, differences in accessibility and availability of care between White people and Black and Latino people increased. In 2018, Black and Latino people were more likely to report delayed care due to lack of transportation and long waiting times at the doctor’s office compared with White people—differences that were not found in 1999.

This study expands the evidence in several ways. First, to the best of our knowledge, this is the first investigation to show worsening racial and ethnic disparities in barriers to timely medical care not related to cost over 20 years. Several studies have reported disparities in some of these measures,^9,10,21^ even in recent years,^8^ but this is the first to quantify how such disparities have changed over an extended period. Second, our evaluation of racial and ethnic trends in 5 specific barriers to timely medical care provides a more comprehensive picture than previous studies, some of which found increasing trends in some of these indicators, but not in all 5, and did not describe trends by race and ethnicity.^12,13^ We found that these barriers increased in prevalence over the 20-year period among Black, Latino/Hispanic, and White people at disparate rates. Third, we describe how the increases in disparities in access to timely medical care occurred mostly among males and were attenuated when stratified by income level. This study is the first, to our knowledge, to evaluate how these racial and ethnic disparities changed by sex and income level.

Our findings have several important health policy implications. First, the increase in prevalence in barriers across racial and ethnic groups in the United States indicates a worsening societal failure to deliver timely medical care. This finding indicates that attempts to improve access to care through improving access to insurance coverage alone may be inadequate. While increasing insurance coverage rates may reduce trends in unmet medical needs due to cost, it is less clear that it can reduce barriers to timely medical care that are not related to cost. In the years after the Affordable Care Act Medicaid expansions, a study found increases in delayed care due to long waiting times and inability to schedule an appointment soon enough, particularly among those with low income.^22^ Similarly, there was no difference in the prevalence of these 2 barriers (long waiting times and appointment availability) by Medicare eligibility status (i.e., those aged <65 years compared with those aged ≥65 years).^23^ This underscores the need for renewed national investments in measuring, tracking, and improving primary care availability and accessibility related to the broader social determinants of health.

Second, the growing racial and ethnic disparities in prevalence in these barriers to timely medical care indicate that the scope of national efforts to eliminate disparities in health care access should be expanded and include societal reforms beyond the health care system. Eliminating disparities in these indicators requires that policy interventions address non-medical barriers to health care access and quality, including education, housing, urban planning, employment, and transportation, which disproportionately impact underserved populations.^24^ Such interventions ought to be implemented in the context of structural racism that accentuates barriers to accessing medical care for minority groups, both within and adjacent to the health care system. Importantly, there is evidence that because of historical segregation of communities of color, Black and Latino individuals are more likely to live in medically underserved areas, to receive worse quality of care, and to visit the emergency department for primary care-treatable conditions.^25-27^ Such disparities are further compounded by transportation barriers.^13^ Thus, there is a need for a multi-sectorial effort to improve spatial accessibility to high-quality primary care clinics and health care professionals for minoritized racial and ethnic groups. Strategies could include addressing differences in distribution of health care facilities, increasing flexibility of care (e.g., implementing urgent clinics that do not result in discontinuity of care), including insurance coverage for non-emergency transportation to medical care, and leveraging digital health technologies for high-quality telehealth consultations that are available and accessible.

Third, there are important implications from the income- and sex-stratified findings. The finding that racial and ethnic disparities were attenuated by lower income serve as an example of the pervasiveness of income inequality in access to health care, even beyond cost-related indicators. Regarding sex, although racial/ethnic disparities among females were mostly static, females had an overall higher prevalence of barriers over the study period compared with males of the same race or ethnicity. As females face structural challenges to accessing sex-specific primary care such as pregnancy and menopause, and gender-sensitive care,^28^ our findings add to the evidence of a need to increase females’ access to primary care during the different stages of their lifetime.

This study has several limitations. First, we measured specific barriers that were consistently ascertained during the study period but were unable to measure other important barriers to timely medical care not related to cost (e.g., language barrier and accessibility to technology accessibility). Second, NHIS data are self-reported, and lacked information on what kind of care was delayed by the measured barriers. Third, there is no information regarding state or rural or urban setting of residence in the publicly available NHIS data, which may influence some of these measures. However, these limitations do not affect the primary findings regarding self-reported barriers over the last 2 decades.

In conclusion, from 1999 to 2018, barriers to timely medical care increased for all 4 racial and ethnic groups included in this study, and there were increasing differences in some of these barriers between groups. Compared with White people, Black and Latino people were more likely to report experiencing these barriers. There is considerable scope to improve availability of medical care and to eliminate these disparities.

## Data Availability

All data used in this study is publicly available and was obtained from the Integrated Public Use Microdata Series Health Surveys (https://nhis.ipums.org/).

