## Supplemental Methods for "Trends in Racial and Ethnic Disparities in Barriers to Timely Medical Care Among US Adults, 1999 to 2018"

### TABLE OF CONTENTS


**Supplemental Methods**

**Supplemental Figure S1. Study Population Flowchart**

**Supplemental Figure S2. Trends in Proportion of Individuals Reporting Delaying Care Because They Couldn't Get Through by Phone by Race and Ethnicity, Stratified by Sex and Income Level**

**Supplemental Figure S3. Trends in Proportion of Individuals Reporting Delaying Care Because They Couldn't Get an Appointment Soon Enough by Race and Ethnicity, Stratified by Sex and Income Level**

**Supplemental Figure S4. Trends in Proportion of Individuals Reporting Delaying Care Because They Had to Wait Too Long to See the Doctor by Race and Ethnicity, Stratified by Sex and Income Level**

**Supplemental Figure S5. Trends in Proportion of Individuals Reporting Delaying Care Because the Doctor's Office Was Not Open When They Could Get There by Race and Ethnicity, Stratified by Sex and Income Level**

**Supplemental Figure S6. Trends in Proportion of Individuals Reporting Delaying Care Because They Lacked Transportation by Race and Ethnicity, Stratified by Sex and Income Level**

**Supplemental Table S1. Study Population Characteristics**

**Supplemental Table S2. Change in the Adjusted Prevalence of Any Barriers to Timely Medical Care from 1999 to 2018, by Race and Ethnicity and Stratified by Sex and Income Level**

**Supplemental Table S3. Change in the Adjusted Proportion of Individuals Reporting Delaying Care Because They Couldn't Get Through the Phone from 1999 to 2018, by Race and Ethnicity and Stratified by Sex and Income Level**

**Supplemental Table S4. Change in the Adjusted Proportion of Individuals Reporting Delaying Care Because They Couldn't Get an Appointment Soon Enough from 1999 to 2018, by Race and Ethnicity and Stratified by Sex and Income Level**

**Supplemental Table S5. Change in the Adjusted Proportion of Individuals Reporting Delaying Care Because They Had to Wait Too Long to See the Doctor from 1999 to 2018, by Race and Ethnicity and Stratified by Sex and Income Level**

**Supplemental Table S6. Change in the Adjusted Proportion of Individuals Reporting Delaying Care Because the Doctor's Office Was Not Open When They Could Get There from 1999 to 2018, by Race and Ethnicity and Stratified by Sex and Income Level**

**Supplemental Table S7. Change in the Adjusted Proportion of Individuals Reporting Delaying Care Because They Lacked Transportation from 1999 to 2018, by Race and Ethnicity and Stratified by Sex and Income Level**

**Supplemental Methods**

*About the National Health Interview Survey*

The National Health Interview Survey (NHIS) consists of a questionnaire divided into 4 cores: Household Composition, Family Core, Sample Child Core, and Sample Adult Core. The Household Composition file collects basic and relationship information about all persons in a household. The Family Core file collects sociodemographic characteristics, basic indicators of health status, activity limitations, injuries, health insurance coverage, and access to and utilization of health care services. From each family, one sample child and one sample adult are randomly selected to gather more in-depth information for the Sample Child Core and Sample Adult Core, respectively.[1]

*Statistical analysis*

In the multivariable logistic regressions to estimate the annual prevalence of each barrier to timely medical care by race/ethnicity, the barrier was the dependent variable, and age and region were the independent variables. Each year's coefficient–combined with the intercept–represented the logit of the adjusted annual proportion of each outcome. Then, we used the results to create estimated annual prevalence by using the inverse logit of each year effect as the annual prevalence, and applying the method of parametric bootstrapping to calculate the standard error (SE) and the confidence interval (CI) for the transformed coefficients.[2]

To estimate the annual low-income prevalence by race and ethnicity, we used the mean annual estimate obtained by separate logistic regressions using a similar approach described in the main analysis, but with each of the multiply imputed low-income variables as the dependent variable and an indicator for each year as the independent variables. Similarly, we used the mean prevalence estimate of each barrier to timely medical care indicator from separate regressions for each of the income groups (low-income and middle/high-income).[3]

In the ordered logistic regression models to estimate the proportion of individuals with 0, 1, 2, 3, or ≥4 specific barriers over the years, the number of barriers was the dependent variable and age, region, and an indicator for each year were the independent variables, obtaining the estimates for each race/ethnicity group separately.

**Supplemental Figure S1.** Study Population Flowchart

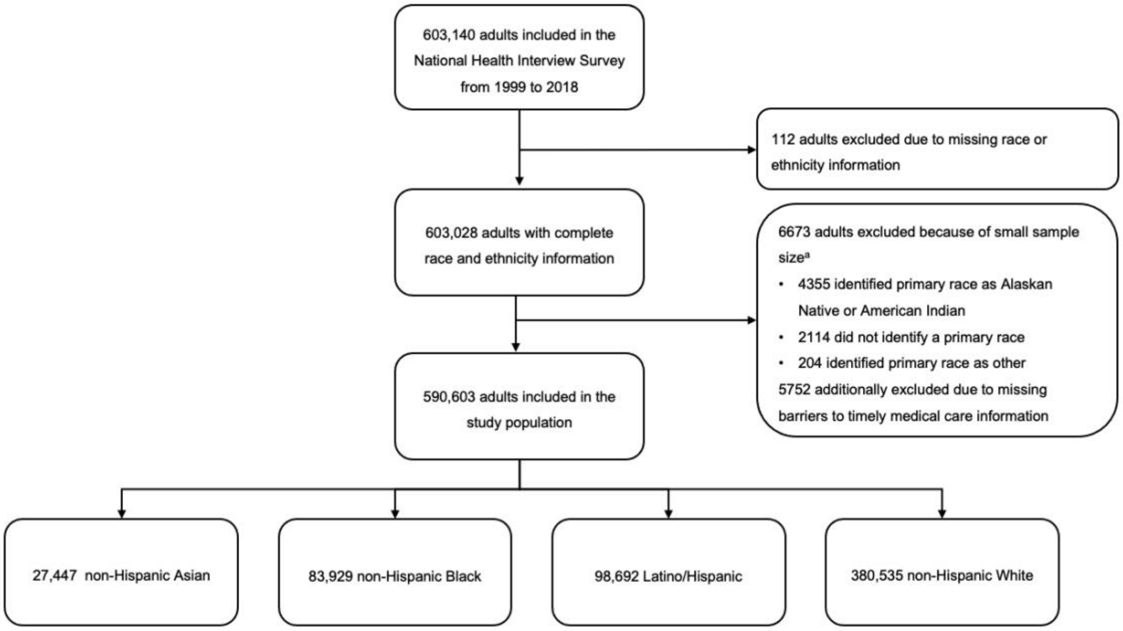


The 4 mutually exclusive racial/ethnic subgroups were created based on the primary race and ethnicity combination.
[a] These excluded individuals also did not identify as Latino/Hispanic.

**Supplemental Figure S2.** Trends in Proportion of Individuals Reporting Delaying Care Because They Couldn't Get Through by Phone by Race and Ethnicity, Stratified by Sex and Income Level

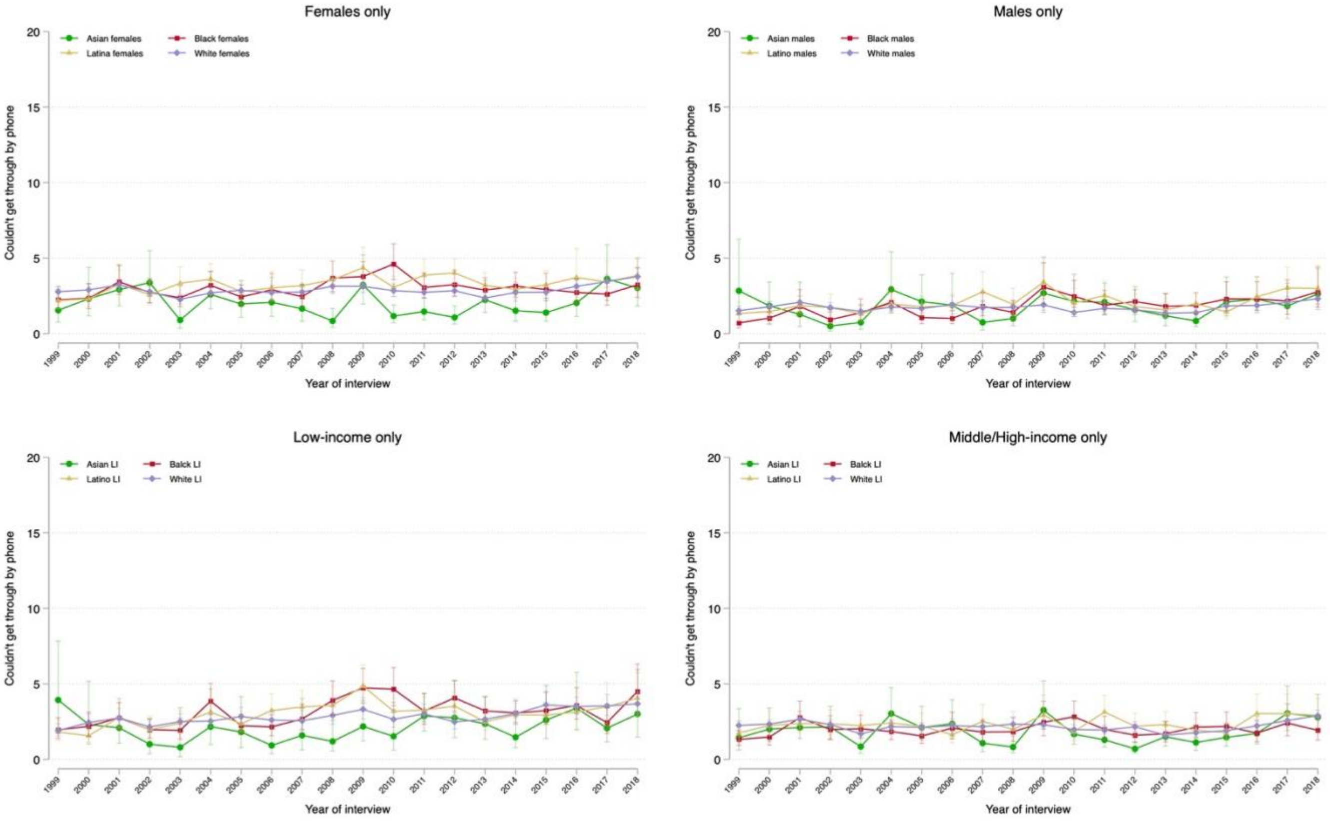

**Supplemental Figure S3.** Trends in Proportion of Individuals Reporting Delaying Care Because They Couldn't Get an Appointment Soon Enough by Race and Ethnicity, Stratified by Sex and Income Level

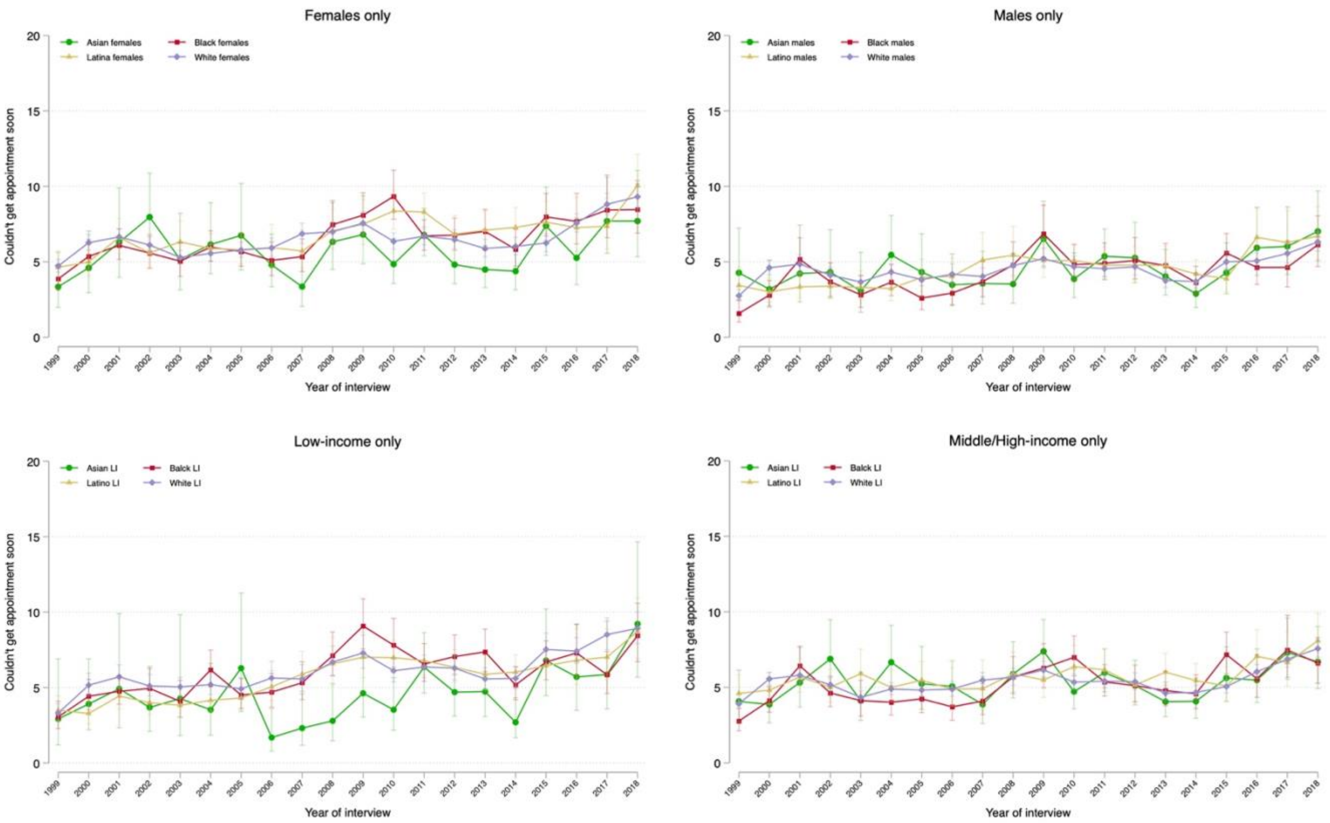

**Supplemental Figure S4.** Trends in Proportion of Individuals Reporting Delaying Care Because They Had to Wait Too Long to See the Doctor by Race and Ethnicity, Stratified by Sex and Income Level

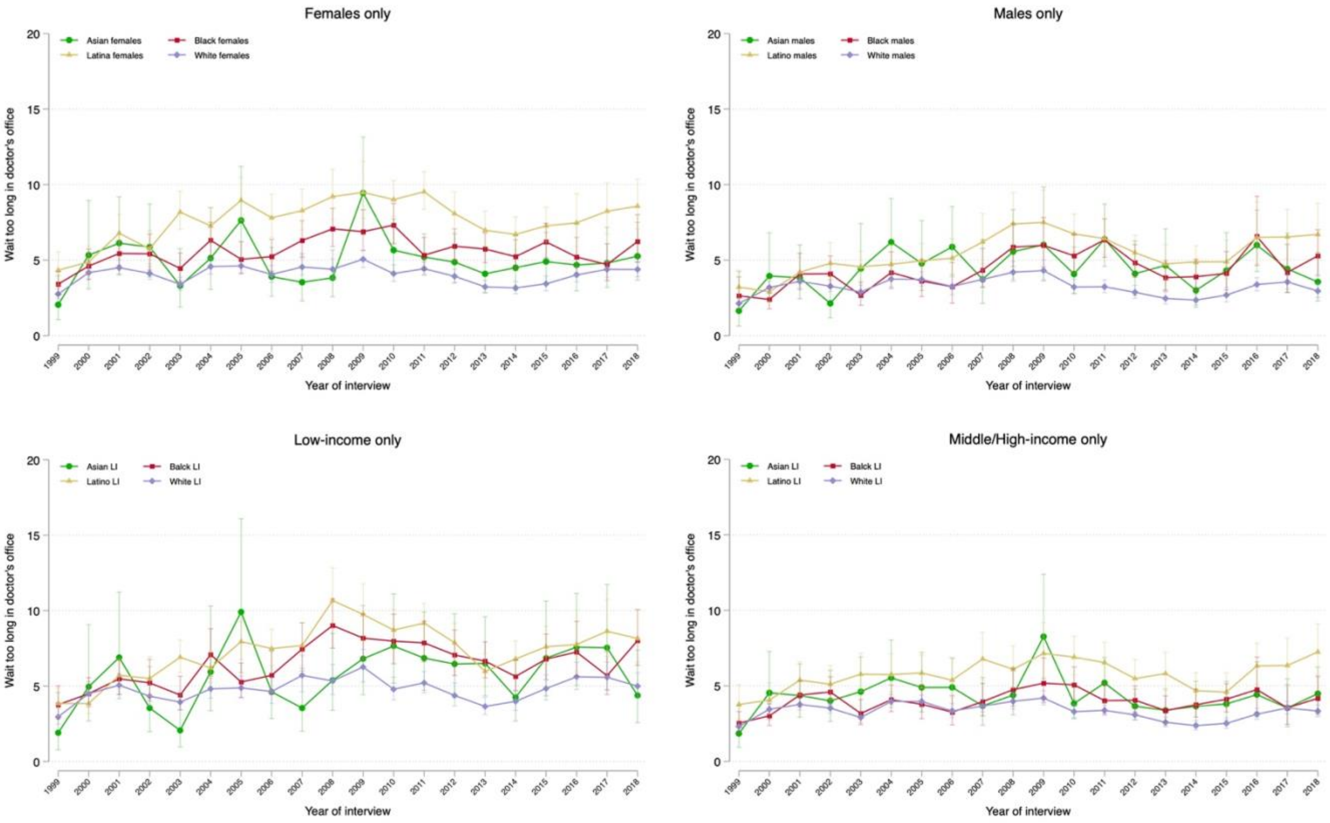

**Supplemental Figure S5.** Trends in Proportion of Individuals Reporting Delaying Care Because the Doctor's Office Was Not Open When They Could Get There by Race and Ethnicity, Stratified by Sex and Income Level

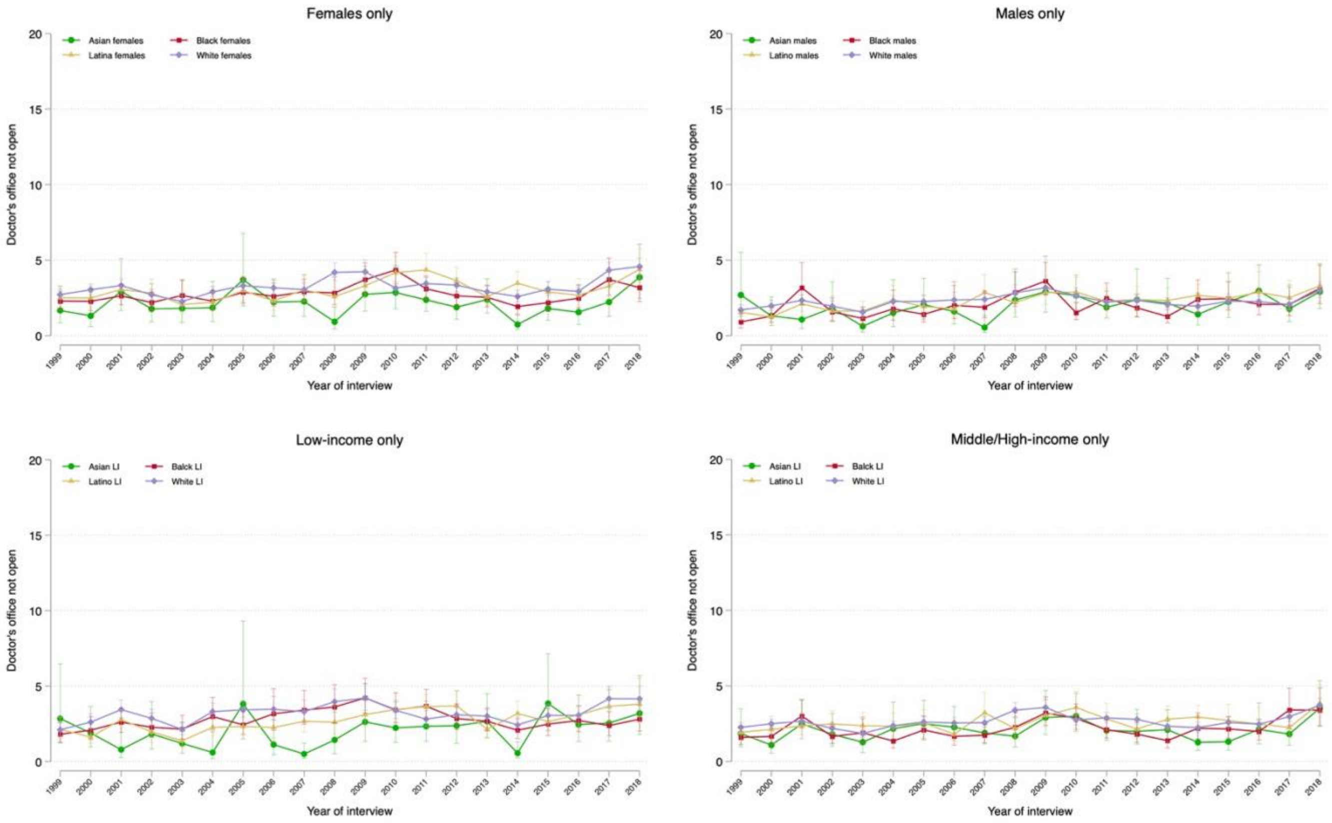

**Supplemental Figure S6.** Trends in Proportion of Individuals Reporting Delaying Care Because They Lacked Transportation by Race and Ethnicity, Stratified by Sex and Income Level

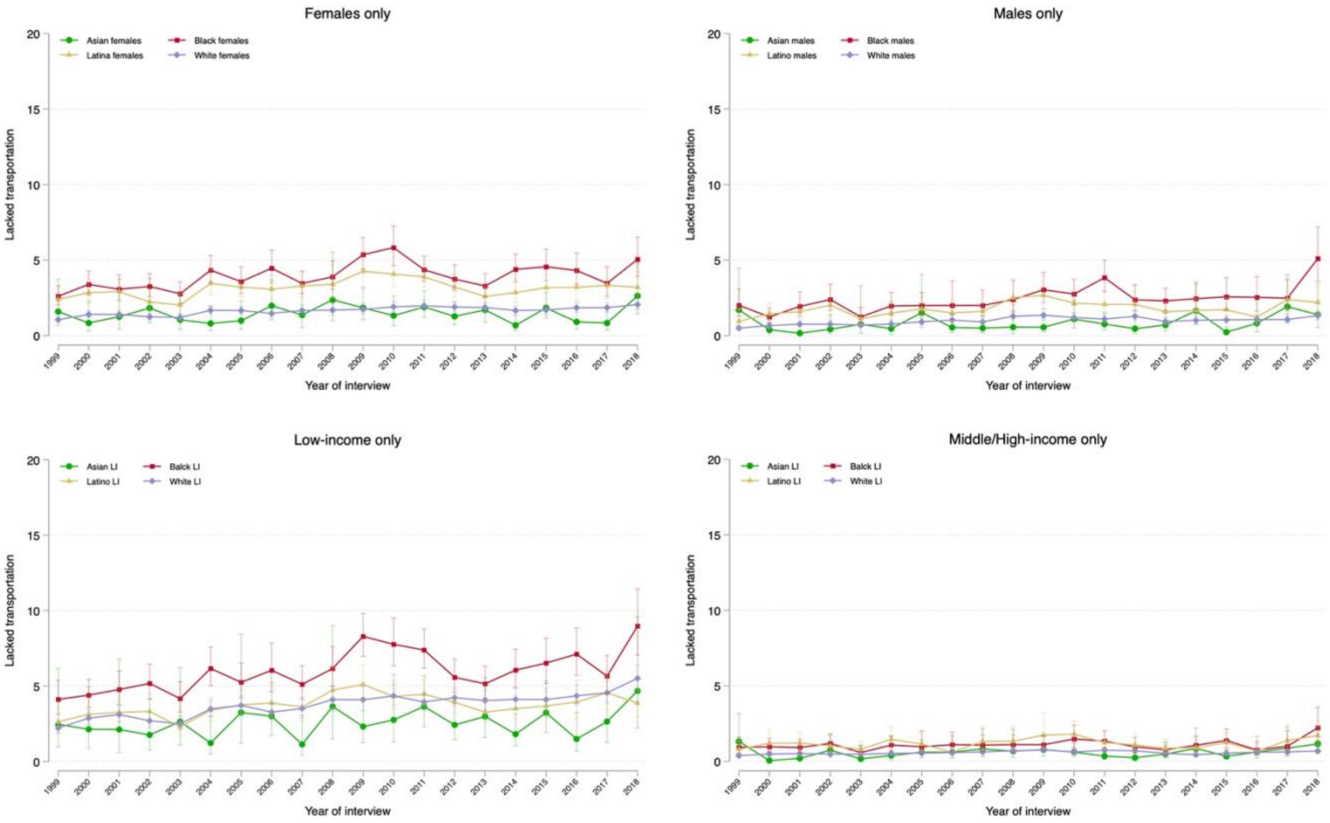

**Supplemental Table S1. Study Population Characteristics by Years**

| | Asian | | | Black | | | Latino/Hispanic | | | White | | |
|---|---|---|---|---|---|---|---|---|---|---|---|---|
| | **1999–2000** | **2008–2009** | **2017–2018** | **1999–2000** | **2008–2009** | **2017–2018** | **1999–2000** | **2008–2009** | **2017–2018** | **1999–2000** | **2008–2009** | **2017–2018** |
| Sample size, n [Total=590,603] | n=1572 | n=2801 | n=2611 | n=8639 | n=7713 | n=5818 | n=10,188 | n=8754 | n=6379 | n=41,503 | n=29,252 | n=36,134 |
| **Age in years** | 38 (28–50) | 41 (30–55) | 43 (32–57) | 40 (29–52) | 42 (29–55) | 43 (30–58) | 37 (27–49) | 37 (28–50) | 39 (28–53) | 44 (33–59) | 47 (33–61) | 50 (34–64) |
| **Age category** | | | | | | | | | | | | |
| 18–39 years | 52.7 (49.5, 55.9) | 46.0 (43.4, 48.7) | 41.9 (39.3, 44.6) | 49.8 (48.3, 51.3) | 45.3 (43.5, 47.1) | 43.7 (41.8, 45.7) | 56.8 (55.3, 58.2) | 54.6 (53.0, 56.2) | 50.1 (48.4, 51.9) | 39.6 (38.9, 40.2) | 34.9 (34.0, 35.8) | 33.3 (32.5, 34.1) |
| 40–64 years | 37.8 (34.7, 40.9) | 40.8 (38.2, 43.5) | 42.5 (40.2, 44.9) | 38.5 (37.2, 39.9) | 42.8 (38.2, 43.5) | 41.0 (39.2, 42.9) | 34.2 (33.1, 35.3) | 37.0 (35.6, 38.4) | 39.1 (37.7, 40.6) | 42.1 (41.5, 42.7) | 45.8 (45.1, 46.6) | 42.3 (41.6, 43.0) |
| ≥65 years | 9.5 (7.9, 11.4) | 13.2 (11.7, 14.8) | 15.5 (13.8, 17.3) | 11.7 (10.9, 12.6) | 12.0 (11.1, 12.9) | 15.3 (14.2, 16.3) | 9.0 (8.1, 10.0) | 8.4 (7.7, 9.1) | 10.8 (9.9, 11.7) | 18.4 (17.8, 18.9) | 19.3 (18.6, 19.9) | 24.4 (23.7, 25.1) |
| **Female** | 51.9 (51.4, 52.5) | 51.7 (51.0, 52.4) | 51.4 (50.7, 52.0) | 55.6 (54.1, 57.1) | 55.4 (53.9, 56.9) | 54.5 (52.8, 56.3) | 50.9 (49.6, 52.1) | 48.7 (47.2, 50.1) | 50.3 (48.7, 52.0) | 51.9 (51.4, 52.5) | 51.7 (51.0, 52.4) | 51.4 (50.7, 52.0) |
| **US citizenship** [n=589,337] | 58.3 (54.7, 61.9) | 69.6 (66.9, 72.2) | 70.8 (68.1, 73.3) | 95.4 (94.7, 96.0) | 95.0 (94.2, 95.6) | 95.0 (93.8, 95.9) | 62.8 (60.7, 64.9) | 62.1 (60.1, 64.0) | 72.4 (70.7, 74.1) | 98.4 (98.2, 98.6) | 98.5 (98.3, 98.7) | 98.5 (98.3, 98.7) |
| **Education level** [n=586,373] | | | | | | | | | | | | |
| Less than high school | 11.4 (9.4, 13.7) | 9.2 (7.9, 10.7) | 8.0 (6.7, 9.6) | 24.4 (23.0, 25.7) | 17.6 (16.3, 19.0) | 14.2 (12.9, 15.7) | 44.5 (42.9, 46.2) | 38.6 (37.0, 40.2) | 27.8 (26.6, 29.7) | 13.4 (12.9, 13.9) | 10.5 (10.0, 11.1) | 7.4 (6.9, 7.9) |
| High school diploma /GED | 18.1 (16.1, 20.3) | 16.2 (14.4, 18.2) | 15.3 (13.4, 17.4) | 31.4 (30.1, 32.8) | 30.7 (29.2, 32.2) | 28.9 (27.3, 30.6) | 24.3 (23.3, 25.4) | 26.6 (25.2, 28.0) | 26.6 (25.1, 28.3) | 31.7 (31.0, 32.4) | 28.2 (27.5, 28.9) | 23.6 (22.9, 24.4) |
| Some college | 24.2 (21.7, 27.0) | 24.4 (22.2, 26.7) | 20.9 (18.9, 23.0) | 29.7 (28.4, 31.0) | 34.0 (32.5, 35.5) | 32.7 (30.9, 34.5) | 21.8 (20.6, 23.1) | 22.5 (21.4, 23.7) | 28.2 (26.6, 29.8) | 29.6 (29.1, 30.1) | 31.6 (30.9, 32.3) | 31.4 (30.6, 32.1) |
| ≥Bachelor's degree | 46.2 (43.2, 49.3) | 50.2 (46.9, 53.5) | 55.9 (52.8, 58.8) | 14.5 (13.4, 15.7) | 17.8 (16.6, 19.0) | 24.2 (22.3, 26.2) | 9.4 (8.5, 10.3) | 12.4 (11.4, 13.4) | 17.4 (16.0, 19.0) | 25.4 (24.7, 26.1) | 29.8 (28.9, 30.7) | 37.6 (36.5, 38.7) |
| **Income <200% of Federal Poverty Limit**[a] | 28.2 (24.0, 32.7) | 27.0 (23.8, 30.4) | 26.3 (22.9, 30.0) | 45.5 (43.2, 47.8) | 45.0 (42.8, 47.3) | 44.5 (41.5, 47.5) | 51.0 (48.9, 53.1) | 50.2 (48.0, 52.3) | 46.7 (44.0, 49.3) | 22.9 (22.1, 23.7) | 23.9 (22.8, 25.0) | 21.3 (20.3, 22.2) |
| **Uninsured at the time of interview** [n=588,490] | 17.3 (15.1, 19.8) | 13.9 (12.1, 15.7) | 6.3 (5.2, 7.6) | 20.2 (19.1, 21.3) | 20.8 (19.5, 22.0) | 11.9 (10.6, 13.4) | 36.1 (34.3, 37.8) | 38.9 (37.0, 40.8) | 24.1 (22.2, 26.0) | 11.0 (10.6, 11.4) | 12.3 (11.8, 12.8) | 6.5 (6.2, 6.9) |
| **Region of residence**[b] | | | | | | | | | | | | |
| Northeast | 21.2 (18.7, 24.0) | 18.6 (16.2, 21.3) | 20.8 (16.8, 25.3) | 17.8 (16.3, 19.4) | 16.4 (14.8, 18.1) | 16.0 (13.5, 18.8) | 15.7 (14.5, 17.0) | 13.3 (11.7, 15.1) | 13.4 (11.2, 16.1) | 20.2 (19.4, 20.9) | 18.2 (17.2, 19.2) | 19.2 (17.5, 21.1) |

| | Asian | | | Black | | | Latino/Hispanic | | | White | | |
|---|---|---|---|---|---|---|---|---|---|---|---|---|
| | **1999–2000** | **2008–2009** | **2017–2018** | **1999–2000** | **2008–2009** | **2017–2018** | **1999–2000** | **2008–2009** | **2017–2018** | **1999–2000** | **2008–2009** | **2017–2018** |
| Midwest | 14.3 (11.6, 17.6) | 15.1 (12.7, 17.8) | 11.9 (9.5, 14.7) | 18.6 (17.1, 20.3) | 19.4 (17.4, 21.5) | 15.1 (12.9, 17.7) | 8.0 (6.9, 9.2) | 9.5 (8.0, 11.2) | 9.4 (7.6, 11.6) | 29.5 (28.6, 30.3) | 28.6 (27.2, 30.0) | 27.4 (25.7, 29.3) |
| South | 19.1 (16.5, 22.0) | 19.4 (17.2, 21.8) | 25.4 (21.4, 29.9) | 56.0 (53.6, 58.3) | 56.1 (53.5, 58.7) | 60.6 (56.8, 64.4) | 35.6 (33.4, 37.9) | 35.4 (33.3, 37.4) | 37.2 (32.6, 42.1) | 33.9 (33.0, 34.9) | 33.8 (32.3, 35.4) | 33.0 (30.8, 35.2) |
| West | 45.4 (41.4, 49.4) | 46.9 (43.6, 50.2) | 42.0 (36.8, 47.2) | 7.6 (6.7, 8.6) | 8.2 (7.3, 9.1) | 8.3 (6.8, 9.9) | 40.8 (38.5, 43.1) | 41.9 (39.5, 44.3) | 39.9 (35.3, 44.8) | 16.5 (15.8, 17.2) | 19.4 (18.3, 20.5) | 20.4 (18.3, 22.7) |
| **Married or living with partner** [n=588,349] | 64.3 (61.2, 67.3) | 64.1 (61.2, 66.9) | 65.0 (62.4, 67.4) | 37.4 (35.9, 38.9) | 35.2 (33.6, 36.8) | 32.7 (31.0, 34.4) | 58.5 (57.3, 59.6) | 54.5 (52.9, 56.1) | 49.3 (47.7, 51.0) | 61.7 (60.9, 62.4) | 58.0 (57.0, 58.9) | 56.4 (55.6, 57.1) |
| **Employment status** [n=589,945] | | | | | | | | | | | | |
| With a job/Working | 65.9 (63.2, 68.5) | 65.1 (62.6, 67.4) | 67.3 (65.0, 69.4) | 64.2 (62.8, 65.6) | 60.8 (59.2, 62.3) | 61.1 (59.0, 63.1) | 66.3 (65.0, 67.5) | 64.1 (62.8, 65.4) | 66.6 (64.7, 68.3) | 65.7 (65.0, 66.4) | 62.6 (61.7, 63.5) | 62.1 (61.3, 62.9) |
| Not in labor force | 31.9 (29.3, 34.7) | 30.3 (28.0, 32.7) | 30.0 (27.7, 32.3) | 31.8 (30.4, 33.6) | 31.2 (29.8, 32.7) | 32.4 (30.5, 34.4) | 31.3 (30.1, 32.5) | 29.0 (27.9, 30.2) | 29.9 (28.0, 31.8) | 32.9 (32.3, 33.6) | 33.3 (32.4, 34.2) | 35.4 (34.7, 36.2) |
| Unemployed | 2.2 (1.5, 3.2) | 4.6 (3.7, 5.7) | 2.8 (2.1, 3.7) | 3.9 (3.4, 4.6) | 8.0 (7.2, 8.9) | 6.6 (5.7, 7.5) | 2.5 (2.1, 2.9) | 6.9 (6.1, 7.7) | 3.6 (3.0, 4.3) | 1.4 (1.2, 1.5) | 4.1 (3.8, 4.4) | 2.5 (2.2, 2.7) |
| **Current smoker** | 14.9 (12.8, 17.2) | 11.0 (9.5, 12.7) | 7.2 (6.1, 8.6) | 23.9 (22.6, 25.2) | 21.3 (20.0, 22.7) | 14.8 (13.5, 16.2) | 18.4 (17.4, 19.3) | 15.1 (14.1, 16.2) | 9.8 (8.9, 10.7) | 24.2 (23.7, 24.8) | 22.1 (21.4, 22.8) | 15.1 (14.5, 15.7) |
| **Obese (BMI ≥30 kg/m$^2$)** | 6.2 (4.9, 7.7) | 8.7 (7.2, 10.6) | 12.0 (10.6, 13.7) | 29.1 (28.0, 30.1) | 37.1 (35.8, 38.5) | 39.5 (37.9, 41.2) | 23.0 (21.9, 24.1) | 31.2 (29.9, 32.4) | 34.0 (32.5, 35.6) | 20.2 (19.7, 20.7) | 26.1 (25.4, 26.8) | 30.3 (29.6, 31.0) |
| **Health conditions** | | | | | | | | | | | | |
| Asthma | 5.6 (4.5, 6.9) | 8.9 (7.6, 10.3) | 8.5 (7.2, 9.9) | 9.0 (8.2, 9.9) | 13.8 (12.8, 13.9) | 15.1 (14.0, 16.2) | 7.1 (6.5, 7.8) | 10.0 (9.2, 10.8) | 11.7 (10.5, 12.9) | 9.2 (8.9, 9.6) | 13.4 (12.9, 13.9 | 13.9 (13.5, 14.4) |
| Cancer | 1.8 (1.1, 2.7) | 3.0 (2.5, 3.7) | 4.2 (3.4, 5.3) | 3.0 (2.6, 3.5) | 3.8 (3.3, 4.3) | 4.8 (4.3, 5.4) | 2.2 (1.8, 2.5) | 2.7 (2.3, 3.2) | 3.4 (2.9, 3.8) | 7.8 (7.6, 8.1) | 10.2 (9.8, 10.6) | 12.4 (12.0, 12.8) |
| COPD | 1.6 (1.0, 2.4) | 1.9 (1.3, 2.7) | 2.0 (1.4, 2.7) | 4.2 (3.8, 4.7) | 4.5 (3.9, 5.1) | 4.4 (3.7, 5.1) | 3.0 (2.6, 3.4) | 2.8 (2.4, 3.3) | 2.8 (2.4, 3.4) | 6.2 (6.0, 6.5) | 6.5 (6.2, 6.9) | 5.3 (5.0, 5.6) |
| Diabetes | 3.9 (3.0, 5.1) | 7.4 (6.2, 8.8) | 9.0 (7.8, 10.4) | 8.2 (7.5, 8.9) | 11.3 (10.5, 12.3) | 11.7 (10.8, 12.7) | 6.4 (5.8, 7.0) | 8.7 (7.9, 9.5) | 10.5 (9.6, 11.4) | 5.2 (5.0, 5.4) | 8.2 (7.8, 8.6) | 9.1 (8.7, 9.5) |
| Heart disease | 4.6 (3.6, 6.0) | 5.0 (4.0, 6.2) | 6.6 (5.6, 7.7) | 8.9 (8.2, 9.7) | 9.8 (8.9, 10.7) | 9.8 (9.0, 10.7) | 6.1 (5.5, 6.7) | 6.1 (5.5, 6.7) | 6.4 (5.7, 7.2) | 12.1 (11.7, 12.4) | 13.8 (13.2, 14.3) | 14.0 (13.6, 14.5) |
| Hypertension | 14.9 (12.9, 17.2) | 22.4 (20.2, 24.7) | 24.2 (22.1, 26.3) | 29.0 (27.7, 30.3) | 35.4 (34.0, 36.9) | 37.6 (35.8, 39.3) | 15.4 (14.5, 16.3) | 19.9 (18.8, 21.1) | 22.3 (20.9, 23.7) | 23.0 (22.5, 23.5) | 30.4 (29.7, 31.1) | 32.7 (31.9, 33.4) |
| Kidney disease | 0.8 (0.4, 1.5) | 1.2 (0.8, 1.7) | 1.8 (1.2, 2.6) | 2.0 (1.6, 2.4) | 2.2 (1.7, 2.7) | 2.6 (2.2, 3.1) | 1.6 (1.3, 1.9) | 1.8 (1.6, 2.0) | 2.0 (1.6, 2.4) | 1.3 (1.2, 1.5) | 1.8 (1.6, 2.0) | 2.3 (2.1, 2.5) |
| Liver disease | 1.1 (0.5, 2.4) | 1.3 (0.9, 2.0) | 1.6 (1.1, 2.4) | 1.0 (0.8, 1.3) | 0.9 (0.7, 1.2) | 1.2 (0.9, 1.5) | 1.0 (0.8, 1.3) | 1.8 (1.5, 2.2) | 2.4 (2.0, 2.9) | 1.0 (0.9, 1.1) | 1.5 (1.3, 1.7) | 1.8 (1.6, 2.0) |

| | Asian | | | Black | | | Latino/Hispanic | | | White | | |
|---|---|---|---|---|---|---|---|---|---|---|---|---|
| | **1999–2000** | **2008–2009** | **2017–2018** | **1999–2000** | **2008–2009** | **2017–2018** | **1999–2000** | **2008–2009** | **2017–2018** | **1999–2000** | **2008–2009** | **2017–2018** |
| Stroke | 0.8 (0.5, 1.2) | 1.2 (0.8, 1.7) | 1.8 (1.4, 2.5) | 2.8 (2.4, 3.2) | 3.3 (2.8, 3.8) | 4.0 (3.5, 4.6) | 1.1 (0.9, 1.4) | 1.6 (1.3, 1.9) | 2.1 (1.7, 2.6) | 2.2 (2.1, 2.3) | 3.0 (2.8, 3.2) | 3.3 (3.1, 3.6) |

Data are presented as % (95% CI) for categorical variables and median (P25–P75) for continuous variables. All percentages are unadjusted and weighted.

[a] Annual family income was categorized relative to the respective year's Federal Poverty Level from the US Census Bureau into middle/high income (≥200%) and low income (<200%). The weighted proportion of individuals with annual income <200% of the Federal Poverty Limit was estimated using multiple imputation.
[b] Based on the Census Bureau-recognized region where the housing unit of the survey participant was located.

Abbreviations: BMI, body mass index; CI, confidence interval; COPD, chronic obstructive pulmonary disease; GED, general equivalency diploma.

**Supplemental Table S2. Change in the Adjusted Prevalence of Any Barriers to Timely Medical Care Access from 1999 to 2018, by Race and Ethnicity and Stratified by Sex and Income Level**

| | **Asian individuals** | **Black individuals** | **Latino/Hispanic individuals** | **White individuals** |
|---|---|---|---|---|
| | Percentage points (95% CI), p value | Percentage points (95% CI), p value | Percentage points (95% CI), p value | Percentage points (95% CI), p value |
| Absolute change in prevalence, 1999–2018 | | | | |
| *Females* | +7.38 (+2.68, +12.08), 0.002 | +7.68 (+4.96, +10.40), <0.001 | +8.52 (+5.83, +11.20), <0.001 | +6.77 (+5.64, +7.91), <0.001 |
| *Males* | +3.89 (-0.63, +8.42), 0.09 | +8.50 (+5.45, +11.54), <0.001 | +7.65 (+4.68, +10.63), <0.001 | +4.93 (+3.93, +5.93), <0.001 |
| *Low-income* | +10.22 (+2.47, +17.97), 0.01 | +10.16 (+6.80, +13.52), <0.001 | +8.95 (+5.91, +11.99), <0.001 | +9.07 (+7.37, +10.77), <0.001 |
| *Middle/high-income* | +4.20 (+0.65, +7.76), 0.02 | +6.42 (+3.97, +8.87), <0.001 | +7.26 (+4.47, +10.04), <0.001 | +5.02 (+4.16, +5.88), <0.001 |
| Difference with White, 1999 | | | | - |
| *Females* | -1.22 (-3.92, +1.48), 0.38 | -0.03 (-1.39, +1.32), 0.96 | +0.78 (-0.70, +2.27), 0.30 | - |
| *Males* | +1.75 (-1.53, +5.04), 0.30 | -0.35 (-1.89, +1.19), 0.66 | +0.99 (-0.69, +2.67), 0.25 | - |
| *Low-income* | -0.65 (-4.63, +3.33), 0.75 | +0.75 (-1.07, +2.57), 0.42 | -0.16 (-1.87, +1.56), 0.86 | - |
| *Middle/high-income* | +0.42 (-2.04, +2.88), 0.74 | -1.16 (-2.48, +0.16), 0.09 | +1.27 (-0.31, +2.86), 0.12 | - |
| Difference with White, 2018 | | | | - |
| *Females* | -0.62 (-4.63, +3.40), 0.76 | +0.87 (-1.74, +3.49), 0.51 | +2.53 (+0.02, +5.03), 0.05 | - |
| *Males* | +0.72 (-2.55, +3.98), 0.67 | +3.22 (+0.41, +6.03), 0.03 | +3.72 (+1.07, +6.37), 0.01 | - |
| *Low-income* | +0.50 (-6.37, +7.36), 0.89 | +1.84 (-1.46, +5.14), 0.27 | -0.28 (-3.31, +2.76), 0.86 | - |
| *Middle/high-income* | -0.39 (-3.09, +2.31), 0.78 | +0.25 (-1.99, +2.48), 0.83 | +3.52 (+1.07, +5.96), 0.01 | - |
| Change in difference with White, 1999–2018 | | | | - |
| *Females* | +0.60 (-4.23, +5.44), 0.81 | +0.91 (-2.04, +3.85), 0.55 | +1.75 (-1.17, +4.66), 0.24 | - |
| *Males* | -1.04 (-5.67, +3.60), 0.66 | +3.57 (+0.36, +6.77), 0.03 | +2.73 (-0.41, +5.86), 0.09 | - |
| *Low-income* | +1.15 (-6.79, +9.08), 0.78 | +1.09 (-2.68, +4.86), 0.57 | -0.12 (-3.60, +3.36), 0.95 | - |
| *Middle/high-income* | -0.81 (-4.47, +2.84), 0.66 | +1.40 (-1.19, +4.00), 0.29 | +2.24 (-0.67, +5.16), 0.13 | - |

Data source is the National Health Interview Survey from years 1999 to 2018. For change in prevalence and change in difference: a positive sign (+) means the prevalence of any barrier to timely medical care (or its difference with White people) increased and a negative sign (-) means it decreased. Estimates were adjusted by age and US region.
Abbreviations: CI, confidence interval

**Supplemental Table S3. Change in the Adjusted Proportion of Individuals Reporting Delaying Care Because They Couldn't Get Through the Phone from 1999 to 2018, by Race and Ethnicity and Stratified by Sex and Income Level**

| | **Asian individuals** | **Black individuals** | **Latino/Hispanic individuals** | **White individuals** |
|---|---|---|---|---|
| | Percentage points (95% CI), p value | Percentage points (95% CI), p value | Percentage points (95% CI), p value | Percentage points (95% CI), p value |
| Absolute change in prevalence, 1999–2018 | | | | |
| *Females* | +1.48 (-0.49, +3.45), 0.14 | +1.00 (-0.19, +2.19), 0.10 | +1.67 (+0.46, +2.88), 0.01 | +1.00 (+0.38, +1.63), 0.002 |
| *Males* | -0.21 (-3.03, +2.61), 0.89 | +2.06 (+0.63, +3.49), 0.01 | +1.66 (+0.23, +3.09), 0.02 | +0.78 (+0.31, +1.26), 0.001 |
| *Low-income* | -0.94 (-4.71, +2.84), 0.63 | +2.54 (+0.80, +4.29), 0.004 | +2.25 (+0.63, +3.86), 0.01 | +1.77 (+0.94, +2.60), <0.001 |
| *Middle/high-income* | +1.37 (-0.49, +3.24), 0.15 | +0.60 (-0.39, +1.59), 0.23 | +1.17 (+0.02, +2.33), 0.05 | +0.64 (+0.19, +1.10), 0.01 |
| Difference with White, 1999 | | | | - |
| *Females* | -1.23 (-2.45, -0.01), 0.05 | -0.54 (-1.25, +0.18), 0.14 | -0.58 (-1.29, +0.13), 0.11 | - |
| *Males* | +1.31 (-1.17, +3.80), 0.30 | -0.81 (-1.36, -0.26), 0.004 | -0.19 (-0.88, +0.50), 0.59 | - |
| *Low-income* | +2.02 (-1.02, +5.07), 0.19 | +0.05 (-0.77, +0.86), 0.91 | -0.10 (-0.90, +0.70), 0.81 | - |
| *Middle/high-income* | -0.82 (-2.24, +0.60), 0.26 | -0.93 (-1.51, -0.35), 0.002 | -0.50 (-1.21, +0.20), 0.16 | - |
| Difference with White, 2018 | | | | - |
| *Females* | -0.75 (-2.42, +0.92), 0.38 | -0.54 (-1.68, +0.60), 0.36 | +0.09 (-1.07, +1.25), 0.88 | - |
| *Males* | +0.32 (-1.09, +1.74), 0.66 | +0.47 (-0.93, +1.87), 0.51 | +0.68 (-0.66, +2.03), 0.32 | - |
| *Low-income* | -0.69 (-3.07, +1.70), 0.57 | +0.82 (-0.94, +2.57), 0.36 | +0.38 (-1.25, +2.00), 0.65 | - |
| *Middle/high-income* | -0.09 (-1.39, +1.21), 0.89 | -0.97 (-1.90, -0.05), 0.04 | +0.03 (-1.00, +1.05), 0.96 | - |
| Change in difference with White, 1999–2018 | | | | - |
| *Females* | +0.48 (-1.59, +2.55), 0.65 | -0.00 (-1.35, +1.34), 0.99 | +0.67 (-0.69, +2.03), 0.34 | - |
| *Males* | -0.99 (-3.85, +1.87), 0.50 | +1.28 (-0.23, +2.79), 0.10 | +0.88 (-0.64, +2.39), 0.26 | - |
| *Low-income* | -2.71 (-6.58, +1.16), 0.17 | +0.77 (-1.16, +2.70), 0.44 | +0.47 (-1.34, +2.29), 0.61 | - |
| *Middle/high-income* | +0.73 (-1.19, +2.65), 0.46 | -0.04 (-1.13, +1.05), 0.94 | +0.53 (-0.72, +1.77), 0.41 | - |

Data source is the National Health Interview Survey from years 1999 to 2018. For change in prevalence and change in difference: a positive sign (+) means the prevalence of this barrier (or its difference with White people) increased and a negative sign (-) means it decreased. Estimates were adjusted by age and US region.
Abbreviations: CI, confidence interval

**Supplemental Table S4. Change in the Adjusted Proportion of Individuals Reporting Delaying Care Because They Couldn't Get an Appointment Soon Enough from 1999 to 2018, by Race and Ethnicity and Stratified by Sex and Income Level**

| | **Asian individuals** | **Black individuals** | **Latino/Hispanic individuals** | **White individuals** |
|---|---|---|---|---|
| | Percentage points (95% CI), p value | Percentage points (95% CI), p value | Percentage points (95% CI), p value | Percentage points (95% CI), p value |
| Absolute change in prevalence, 1999–2018 | | | | |
| *Females* | +4.35 (+0.94, +7.76), 0.01 | +4.59 (+2.63, +6.55), <0.001 | +5.42 (+3.28, +7.56), <0.001 | +4.59 (+3.65, +5.53), <0.001 |
| *Males* | +2.74 (-0.57, +6.06), 0.11 | +4.55 (+2.71, +6.40), <0.001 | +3.26 (+1.21, +5.32), 0.002 | +3.57 (+2.82, +4.31), <0.001 |
| *Low-income* | +6.30 (+0.87,11.73), 0.02 | +5.38 (+3.19, +7.58), <0.001 | +5.24 (+3.07, +7.41), <0.001 | +5.62 (+4.39, +6.85), <0.001 |
| *Middle/high-income* | +2.62 (-0.07, +5.32), 0.06 | +3.84 (+2.12, +5.57), <0.001 | +3.48 (+1.47, +5.48), <0.001 | +3.66 (+2.97, +4.34), <0.001 |
| Difference with White, 1999 | | | | - |
| *Females* | -1.37 (-3.26, +0.51), 0.15 | -0.85 (-1.81, +0.11), 0.08 | -0.09 (-1.19, +1.01), 0.87 | - |
| *Males* | +1.52 (-0.88, +3.93), 0.22 | -1.18 (-2.00, -0.35), 0.01 | +0.69 (-0.44, +1.82), 0.23 | - |
| *Low-income* | -0.39 (-3.40, +2.63), 0.80 | -0.25 (-1.35, +0.84), 0.65 | +0.19 (-0.86, +1.24), 0.73 | - |
| *Middle/high-income* | +0.16 (-1.60, +1.91), 0.86 | -1.14 (-1.98, -0.31), 0.01 | +0.69 (-0.46, +1.85), 0.24 | - |
| Difference with White, 2018 | | | | - |
| *Females* | -1.61 (-4.60, +1.38), 0.29 | -0.85 (-2.80, +1.10), 0.39 | +0.74 (-1.32, +2.80), 0.48 | - |
| *Males* | +0.70 (-1.71, +3.10), 0.57 | -0.19 (-2.00, +1.63), 0.84 | +0.38 (-1.48, +2.25), 0.69 | - |
| *Low-income* | +0.30 (-4.38, +4.97), 0.90 | -0.49 (-2.75, +1.77), 0.67 | -0.19 (-2.45, +2.07), 0.87 | - |
| *Middle/high-income* | -0.88 (-3.03, +1.28), 0.42 | -0.96 (-2.61, +0.70), 0.26 | +0.51 (-1.26, +2.29), 0.57 | - |
| Change in difference with White, 1999–2018 | | | | - |
| *Females* | -0.24 (-3.77, +3.30), 0.90 | +0.00 (-2.17, +2.18), 0.99 | +0.83 (-1.50, +3.17), 0.49 | - |
| *Males* | -0.82 (-4.22, +2.58), 0.64 | +0.99 (-1.00, +2.98), 0.33 | -0.30 (-2.49, +1.88), 0.79 | - |
| *Low-income* | +0.68 (-4.88, +6.24), 0.81 | -0.23 (-2.75, +2.28), 0.86 | -0.38 (-2.87, +2.11), 0.77 | - |
| *Middle/high-income* | -1.03 (-3.81, +1.75), 0.47 | +0.19 (-1.67, +2.04), 0.84 | -0.18 (-2.30, +1.94), 0.87 | - |

Data source is the National Health Interview Survey from years 1999 to 2018. For change in prevalence and change in difference: a positive sign (+) means the prevalence of this barrier to timely medical care (or its difference with White people) increased and a negative sign (-) means it decreased. Estimates were adjusted by age and US region.
Abbreviations: CI, confidence interval

**Supplemental Table S5. Change in the Adjusted Proportion of Individuals Reporting Delaying Care Because They Had to Wait Too Long to See the Doctor from 1999 to 2018, by Race and Ethnicity and Stratified by Sex and Income Level**

| | **Asian individuals** | **Black individuals** | **Latino/Hispanic individuals** | **White individuals** |
|---|---|---|---|---|
| | Percentage points (95% CI), p value | Percentage points (95% CI), p value | Percentage points (95% CI), p value | Percentage points (95% CI), p value |
| Absolute change in prevalence, 1999–2018 | | | | |
| *Females* | +3.21 (+0.79, +5.63), 0.01 | +2.82 (+1.05, +4.58), 0.002 | +4.24 (+2.27, +6.21), <0.001 | +1.63 (+1.03, +2.22), <0.001 |
| *Males* | +1.92 (-0.54, +4.37), 0.13 | +2.65 (+0.79, +4.51), 0.01 | +3.48 (+1.39, +5.58), 0.001 | +0.82 (+0.23, +1.41), 0.01 |
| *Low-income* | +2.47 (-0.69, +5.63), 0.13 | +4.25 (+2.05, +6.45), <0.001 | +4.31 (+2.33, +6.30), <0.001 | +2.03 (+1.00, +3.06), <0.001 |
| *Middle/high-income* | +2.63 (+0.57, +4.70), 0.01 | +1.64 (+0.15, +3.13), 0.03 | +3.47 (+1.43, +5.52), <0.001 | +1.02 (+0.55, +1.50), <0.001 |
| Difference with White, 1999 | | | | - |
| *Females* | -0.71 (-2.20, +0.79), 0.35 | +0.66 (-0.16, +1.48), 0.12 | +1.57 (+0.42, +2.72), 0.01 | - |
| *Males* | -0.49 (-2.37, +1.40), 0.61 | +0.51 (-0.61, +1.62), 0.37 | +1.09 (+0.03, +2.15), 0.04 | - |
| *Low-income* | -1.06 (-3.13, +1.02), 0.32 | +0.80 (-0.46, +2.06), 0.22 | +0.88 (-0.24, +2.00), 0.125 | - |
| *Middle/high-income* | -0.46 (-1.89, +0.96), 0.53 | +0.23 (-0.53, +0.98), 0.55 | +1.46 (+0.29, +2.63), 0.02 | - |
| Difference with White, 2018 | | | | - |
| *Females* | +0.88 (-1.11, +2.87), 0.39 | +1.85 (+0.18, +3.52), 0.03 | +4.19 (+2.48, +5.90), <0.001 | - |
| *Males* | +0.60 (-1.08, +2.29), 0.48 | +2.33 (+0.73, +3.94), 0.004 | +3.75 (+1.84, +5.65), <0.001 | - |
| *Low-income* | -0.61 (-3.21, +1.98), 0.64 | +3.02 (+0.94, +5.10), 0.004 | +3.16 (+1.23, +5.10), 0.001 | - |
| *Middle/high-income* | +1.15 (-0.42, +2.72), 0.15 | +0.85 (-0.52, +2.21), 0.23 | +3.91 (+2.17, +5.66), <0.001 | - |
| Change in difference with White, 1999–2018 | | | | - |
| *Females* | +1.59 (-0.90, +4.08), 0.21 | +1.19 (-0.67, +3.05), 0.21 | +2.62 (+0.56, +4.68), 0.01 | - |
| *Males* | +1.09 (-1.44, +3.62), 0.40 | +1.82 (-0.13, +3.78), 0.07 | +2.66 (+0.48, +4.83), 0.02 | - |
| *Low-income* | +0.45 (-2.88, +3.77), 0.79 | +2.22 (-0.21, +4.65), 0.07 | +2.28 (+0.05, +4.52), 0.05 | - |
| *Middle/high-income* | +1.61 (-0.51, +3.74), 0.14 | +0.62 (-0.95, +2.18), 0.44 | +2.45 (+0.35, +4.55), 0.02 | - |

Data source is the National Health Interview Survey from years 1999 to 2018. For change in prevalence and change in difference: a positive sign (+) means the prevalence of this barrier to timely medical care (or its difference with White people) increased and a negative sign (-) means it decreased. Estimates were adjusted by age and US region.
Abbreviations: CI, confidence interval

**Supplemental Table S6. Change in the Adjusted Proportion of Individuals Reporting Delaying Care Because the Doctor's Office Was Not Open When They Could Get There from 1999 to 2018, by Race and Ethnicity and Stratified by Sex and Income Level**

| | Asian individuals | Black individuals | Latino/Hispanic individuals | White individuals |
|---|---|---|---|---|
| | Percentage points (95% CI), p value | Percentage points (95% CI), p value | Percentage points (95% CI), p value | Percentage points (95% CI), p value |
| Absolute change in prevalence, 1999–2018 | | | | |
| *Females* | +2.20 (+0.01, +4.38), 0.05 | +0.90 (-0.39, +2.18), 0.17 | +1.87 (+0.51, +3.24), 0.01 | +1.86 (+1.22, +2.50), <0.001 |
| *Males* | +0.22 (-2.40, +2.84), 0.87 | +2.23 (+0.81, +3.64), 0.002 | +1.75 (+0.33, +3.18), 0.02 | +1.31 (+0.70, +1.92), <0.001 |
| *Low-income* | +0.34 (-3.01, +3.70), 0.84 | +1.00 (-0.20, +2.20), 0.10 | +1.67 (+0.05, +3.29), 0.04 | +2.04 (+1.11, +2.98), <0.001 |
| *Middle/high-income* | +1.68 (-0.31, +3.67), 0.10 | +1.80 (+0.37, +3.23), 0.01 | +1.93 (+0.69, +3.16), 0.002 | +1.47 (+0.95, +1.99), <0.001 |
| Difference with White, 1999 | | | | - |
| *Females* | -1.05 (-2.33, +0.23), 0.11 | -0.43 (-1.12, +0.25), 0.22 | -0.20 (-0.95, +0.55), 0.61 | - |
| *Males* | +0.99 (-1.18, +3.15), 0.37 | -0.79 (-1.46, -0.13), 0.02 | -0.16 (-0.92, +0.61), 0.69 | - |
| *Low-income* | +0.73 (-2.02, +3.49), 0.60 | -0.31 (-1.10, +0.49), 0.45 | +0.02 (-0.82, +0.86), 0.97 | - |
| *Middle/high-income* | -0.41 (-1.73, +0.91), 0.54 | -0.66 (-1.32, +0.00), 0.05 | -0.32 (-1.01, +0.36), 0.36 | - |
| Difference with White, 2018 | | | | - |
| *Females* | -0.71 (-2.59, +1.17), 0.46 | -1.40 (-2.66, -0.13), 0.03 | -0.19 (-1.49, +1.12), 0.78 | - |
| *Males* | -0.10 (-1.69, +1.49), 0.90 | +0.12 (-1.27, +1.51), 0.86 | +0.29 (-1.06, +1.64), 0.68 | - |
| *Low-income* | -0.97 (-3.10, +1.17), 0.38 | -1.35 (-2.64, -0.05), 0.04 | -0.36 (-2.03, +1.31), 0.68 | - |
| *Middle/high-income* | -0.19 (-1.77, +1.39), 0.81 | -0.33 (-1.69, +1.04), 0.64 | +0.14 (-1.01, +1.29), 0.82 | - |
| Change in difference with White, 1999–2018 | | | | - |
| *Females* | +0.34 (-1.94, +2.61), 0.77 | -0.96 (-2.40, +0.47), 0.19 | +0.01 (-1.50, +1.52), 0.99 | - |
| *Males* | -1.09 (-3.78, +1.60), 0.43 | +0.92 (-0.62, +2.45), 0.24 | +0.44 (-1.11, +2.00), 0.58 | - |
| *Low-income* | -1.70 (-5.19, +1.78), 0.34 | -1.04 (-2.56, +0.48), 0.18 | -0.37 (-2.24, +1.50), 0.70 | - |
| *Middle/high-income* | +0.22 (-1.84, +2.27), 0.84 | +0.33 (-1.19, +1.85), 0.67 | +0.46 (-0.88, +1.80), 0.50 | - |

Data source is the National Health Interview Survey from years 1999 to 2018. For change in prevalence and change in difference: a positive sign (+) means the prevalence of this barrier to timely medical care (or its difference with White people) increased and a negative sign (-) means it decreased. Estimates were adjusted by age and US region.
Abbreviations: CI, confidence interval

**Supplemental Table S7. Change in the Adjusted Proportion of Individuals Reporting Delaying Care Because They Lacked Transportation from 1999 to 2018, by Race and Ethnicity and Stratified by Sex and Income Level**

| | **Asian individuals** | **Black individuals** | **Latino/Hispanic individuals** | **White individuals** |
|---|---|---|---|---|
| | Percentage points (95% CI), p value | Percentage points (95% CI), p value | Percentage points (95% CI), p value | Percentage points (95% CI), p value |
| Absolute change in prevalence, 1999–2018 | | | | |
| *Females* | +1.04 (-1.26, +3.34), 0.37 | +2.44 (+0.98, +3.90), 0.001 | +0.82 (-0.29, +1.92), 0.15 | +1.00 (+0.60, +1.41), <0.001 |
| *Males* | -0.32 (-2.77, +2.14), 0.80 | +3.09 (+1.05, +5.13), 0.003 | +1.25 (+0.17, +2.33), 0.02 | +0.83 (+0.50, +1.16), <0.001 |
| *Low-income* | +2.21 (-2.40, +6.83), 0.35 | +4.86 (+2.37, +7.35), <0.001 | +1.24 (-0.09, +2.57), 0.07 | +3.29 (+2.37, +4.22), <0.001 |
| *Middle/high-income* | -0.16 (-1.78, +1.45), 0.84 | +1.29 (+0.01, +2.56), 0.05 | +0.91 (-0.01, +1.83), 0.05 | +0.31 (+0.09, +0.53), 0.01 |
| Difference with White, 1999 | | | | - |
| *Females* | +0.54 (-1.02, +2.10), 0.50 | +1.56 (+0.90, +2.21), <0.001 | +1.34 (+0.63, +2.05), <0.001 | - |
| *Males* | +1.21 (-0.73, +3.15), 0.22 | +1.51 (+0.58, +2.45), 0.001 | +0.45 (-0.20, +1.11), 0.17 | - |
| *Low-income* | +0.24 (-2.45, +2.94), 0.86 | +1.90 (+0.69, +3.11), 0.002 | +0.41 (-0.49, +1.32), 0.37 | - |
| *Middle/high-income* | +0.94 (-0.43, +2.31), 0.18 | +0.55 (-0.04, +1.14), 0.07 | +0.42 (-0.08, +0.92), 0.10 | - |
| Difference with White, 2018 | | | | - |
| *Females* | +0.57 (-1.16, +2.31), 0.52 | +2.99 (+1.63, +4.36), <0.001 | +1.15 (+0.21, +2.09), 0.02 | - |
| *Males* | +0.06 (-1.49, +1.61), 0.94 | +3.78 (+1.93, +5.62), <0.001 | +0.87 (-0.05, +1.79), 0.06 | - |
| *Low-income* | -0.84 (-4.69, +3.02), 0.67 | +3.47 (+1.10, +5.83), 0.004 | -1.64 (-2.98, -0.30), 0.02 | - |
| *Middle/high-income* | +0.47 (-0.42, +1.35), 0.30 | +1.53 (+0.38, +2.68), 0.01 | +1.03 (+0.23, +1.83), 0.02 | - |
| Change in difference with White, 1999–2018 | | | | - |
| *Females* | +0.04 (-2.29, +2.37), 0.98 | +1.44 (-0.07, +2.95), 0.06 | -0.19 (-1.36, +0.99), 0.76 | - |
| *Males* | -1.15 (-3.63, +1.33), 0.37 | +2.26 (+0.19, +4.33), 0.03 | +0.42 (-0.71, +1.55), 0.47 | - |
| *Low-income* | -1.08 (-5.79, +3.63), 0.65 | +1.57 (-1.09, +4.23), 0.25 | -2.05 (-3.67, -0.43), 0.01 | - |
| *Middle/high-income* | -0.47 (-2.10, +1.16), 0.57 | +0.98 (-0.31, +2.27), 0.14 | +0.61 (-0.34, +1.55), 0.21 | - |

Data source is the National Health Interview Survey from years 1999 to 2018. For change in prevalence and change in difference: a positive sign (+) means the prevalence of this barrier to timely medical care (or its difference with White people) increased and a negative sign (-) means it decreased. Estimates were adjusted by age and US region.
Abbreviations: CI, confidence interval
